# Three month FVC change: a trial endpoint for IPF based on individual participant data meta-analysis

**DOI:** 10.1101/2021.09.13.21263381

**Authors:** Fasihul A Khan, Iain Stewart, Samuel Moss, Laura Fabbri, Karen A. Robinson, Simon Johnson, R Gisli Jenkins

## Abstract

**Rationale:** Novel therapies for idiopathic pulmonary fibrosis (IPF) are in development, but there remains uncertainty about the optimal trial endpoint. An earlier endpoint would enable assessment of a greater number of therapies in adaptive trial designs.

**Objectives:** Individual participant data (IPD) from placebo arms of interventional trials were sought to determine whether short-term changes in forced vital capacity (FVC), gas transfer for carbon monoxide (DL_CO_) and six-minute walk distance (6MWD) could act as surrogate endpoints to accelerate early-phase trials in IPF.

**Methods:** Electronic databases were searched on 1^st^ December 2020, and IPD were sought and meta-analysed. The primary outcome was overall mortality according to baseline and/or three-month change in either FVC, DL_CO_ or 6MWD, with a secondary outcome of disease progression at 12 months, adjusted for age, sex, smoking status and baseline physiology.

**Measurements and main results:** IPD was available for 10/23 eligible studies totalling 1819 participants. Baseline and three-month change in all physiological variables were independently associated with disease outcomes. A 2.5% relative decline in FVC over three months was associated with mortality (adjusted hazard ratio 1.14, 95%CI 1.06;1.24, I^2^ = 59.9%) and disease progression (adjusted odds ratio 1.29; 95%CI 1.18;1.40, I^2^=67%). Optimal thresholds for three-month change in FVC for distinguishing disease outcomes were identified.

**Conclusions:** IPD meta-analysis of trial placebo arms demonstrated three-month change in physiological variables, particularly FVC, were associated with mortality and disease progression among individuals with untreated IPF. FVC change over three months may hold potential as a surrogate endpoint in IPF interventional adaptive trials.

## INTRODUCTION

Idiopathic pulmonary fibrosis (IPF) is a progressive fibrotic lung disease of unknown aetiology characterised by dyspnoea and reduced exercise tolerance. Prognosis is poor, with a median survival of three years.(1) However, there is substantial heterogeneity in disease trajectory, ranging from stable disease to rapid progression associated with significant lung function deterioration, respiratory failure and death.(2)

In recent years, several interventional randomised controlled trials (RCT) in IPF have been performed but defining the optimal primary endpoint on which to design such clinical trials remains the subject of ongoing debate. Mortality may be the ideal primary endpoint but is considered impractical due to the sample sizes required, and therefore surrogate markers including change in FVC over 12 months are accepted by drug regulators as primary endpoints in IPF clinical trials.(3, 4) However, in a progressive disease with poor survival, an endpoint measured at 12 months is unsatisfactory and leads to delayed completion of clinical trials. An earlier endpoint would enable assessment of a greater number of ever-increasing potential therapies and allow adaptive trial designs to be considered. Therefore, we sought to determine whether short term changes (i.e., 3 months) in commonly measured physiological measurements, forced vital capacity (FVC), gas transfer for carbon monoxide (DL_CO_) and six-minute walk distance (6MWD), would predict mortality and thus streamline trials, including allowing for adaptive trial designs in IPF.

Placebo arms of interventional trials offer an invaluable resource to investigate whether commonly measured characteristics can accurately predict disease outcomes. They also enable exploration of the natural trajectory of rare disease as participants are naïve to novel therapeutics, which minimises confounding. We combined data from clinical trial placebo arms using individual patient data (IPD) meta-analysis, considered a gold-standard of systematic review, to explore short term change in physiology (FVC, DL_CO_ and 6MWD) as surrogate markers for disease outcomes, which can then be used as endpoints in future IPF clinical trials.

## METHODS

The systematic review and IPD meta-analysis were conducted in accordance with a pre-specified protocol (PROSPERO registration number: CRD42020164935) and has been reported using PRISMA-IPD (Preferred Reporting Items for Systematic Review and Meta-Analyses of Individual Participant Data) guidelines.(5)

### Search strategy and study selection

Electronic database searches were performed in MEDLINE (1946 to latest), Embase (1974 to latest), Google Scholar, ClinicalTrials.gov and preprint servers including medRxiv and bioRxiv, with the last search on 1^st^ December 2020. Keywords were applied according to PICOS search terms summarised as: Population, idiopathic pulmonary fibrosis; Intervention, physiological exposure (FVC, DL_CO_, 6MWD); Outcome, overall mortality, or disease progression; Study, human clinical trials reported in English (Figure E1). Searches were supplemented by hand-searching reference lists of included studies. Two reviewers (FK and LF) independently screened titles and abstracts, and then full text of retrieved articles.

Studies were eligible for inclusion if they included baseline and/or three-month change in either FVC, DL_CO_ or 6MWD. Non-randomised, retrospective studies, conference abstracts, studies investigating non-IPF interstitial lung disease and studies with a sample size fewer than 30 participants were excluded. To minimise the effect of anti-fibrotic treatment, we restricted inclusion criteria to placebo arms of RCTs reporting outcomes in patients aged over 18 with anti-fibrotic naïve IPF, diagnosed according to contemporaneous consensus guidelines.(6-8) Duplicate articles reporting outcomes from the same cohort were restricted to the original trial publication.

The primary outcome was overall mortality. Secondary outcomes included disease progression at 12 months defined as ≥10% FVC decline or death, as well as absolute change in FVC (ml) between baseline and 12 months.

### Data extraction and risk of bias assessment

IPD were sought from corresponding authors using secure and encrypted electronic mail communication, with a maximum of three reminders sent monthly. Data from sponsored clinical studies were requested through online secure data portals, with signed data-sharing agreements.(9-11) Requested data included participant demographics (age, sex, smoking status), baseline and three-month physiology data, and outcome data including 12-month lung function and overall mortality. Where IPD were unavailable, studies were ineligible for inclusion, but demographics were tabulated to address selection bias. Data extraction was carried out by one reviewer and verified for consistency and accuracy by a second reviewer.

Risk of bias assessment was carried out independently in duplicate using a modified version of the Quality in Prognostic Studies (QUIPS) tool.(12) No specific tools are available for assessing the risk of bias in placebo arms of RCTs, and therefore we assessed bias by considering placebo arms as observational cohorts. The modified QUIPS tool assessed the risk of bias across five domains: study participation, study attrition, prognostic factor measurement, outcome measurement and study confounding. All studies were included in the review irrespective of their risk of bias rating. The GRADE (Grading of Recommendations, Assessment, Development and Evaluations) framework was applied to assess the certainty of evidence for each of the outcomes. (13)

### Statistical analysis

Study and baseline participant characteristics were described in summary tables. Hazard ratios (HR) for associations with overall mortality, and odds ratio (OR) for association with disease progression, were estimated using IPD meta-analysis with random effects and inverse variance weighting in a two-step design to support inclusion of studies across secure servers.(14) FVC predicted reference values were standardised using Global Lung Initiative (GLI) equations and applied to all IPD where possible.(15) Estimates were adjusted for covariates identified *a priori* including age, sex, smoking history and baseline FVC (litres). Participants with missing data were excluded using *listwise deletion*. Disease progression was dichotomised and defined as 10% relative decline in FVC or death within 12 months of baseline.

Baseline continuous associations with outcomes were estimated per 5% decline in %predicted FVC and DLCO, and 50m decline in 6MWD. A baseline cut point of %predicted FVC at 80% was also used to define subgroups based on frequently used criteria in anti-fibrotic management.(16) The association of continuous physiological variable change over three months with outcomes were estimated using 2.5% relative percent change from baseline %predicted FVC and DLCO, and 20m decline in 6MWD. Median relative percent change was used to assess mean difference in absolute FVC (ml) between baseline and 12 months based on subgroups. Optimal cut-points based on sensitivities and specificities of three-month FVC change in predicting disease progression and death were estimated individually for each study with the empirical Liu method and bootstrapping to derive robust confidence intervals.(17) Empirical cut-points were included in meta-analysis to estimate an overall threshold for three-month FVC change and used to estimate association of outcome in those with decline greater than threshold compared to those below. IPD meta-analysis of the area under the receiver operator characteristics curve (AUROC) was used to assess discriminative performance of overall estimated relative 3-month FVC decline cut-points.

Inspection of visual plots and the I^2^ statistic was used to evaluate the level of statistical heterogeneity. Meta-regression was conducted where appropriate to explore heterogeneity according to various study level factors, including permitted concomitant steroid use, the inclusion of participants diagnosed within five years, and the inclusion of severe disease (FVC<50% predicted). Publication bias was considered using visual assessment of funnel plot asymmetry and Egger’s test where possible.(18) All statistical analyses were performed using Stata 16 (Statacort, Texas US).

## RESULTS

Searches of the electronic databases carried out on 1^st^ December 2020 yielded 271 articles, with a further 5 articles identified through ClinicalTrials.gov. After screening of abstract and titles, and full text review, 23 studies with a total of 2958 participants were eligible for inclusion (Figure 1). Corresponding authors and study sponsors were contacted for IPD, which were made available for 10 studies reporting outcomes from 12 individual placebo cohorts totalling 1819 participants. IPD could not be attained from the remaining 13 studies (Table E4). All included studies were multi-centre RCTs published between 2005 and 2014. In 75% (8/12) of studies, an FVC ≥ 50% predicted, and a minimum DL_CO_ of at least either 30% or 35% predicted, were part of the inclusion criteria. In 7 studies, participants in placebo arms were allowed concomitant prednisolone, typically up to a maximum daily dose of 15-20mg. (Table 1).

**Table 1.**
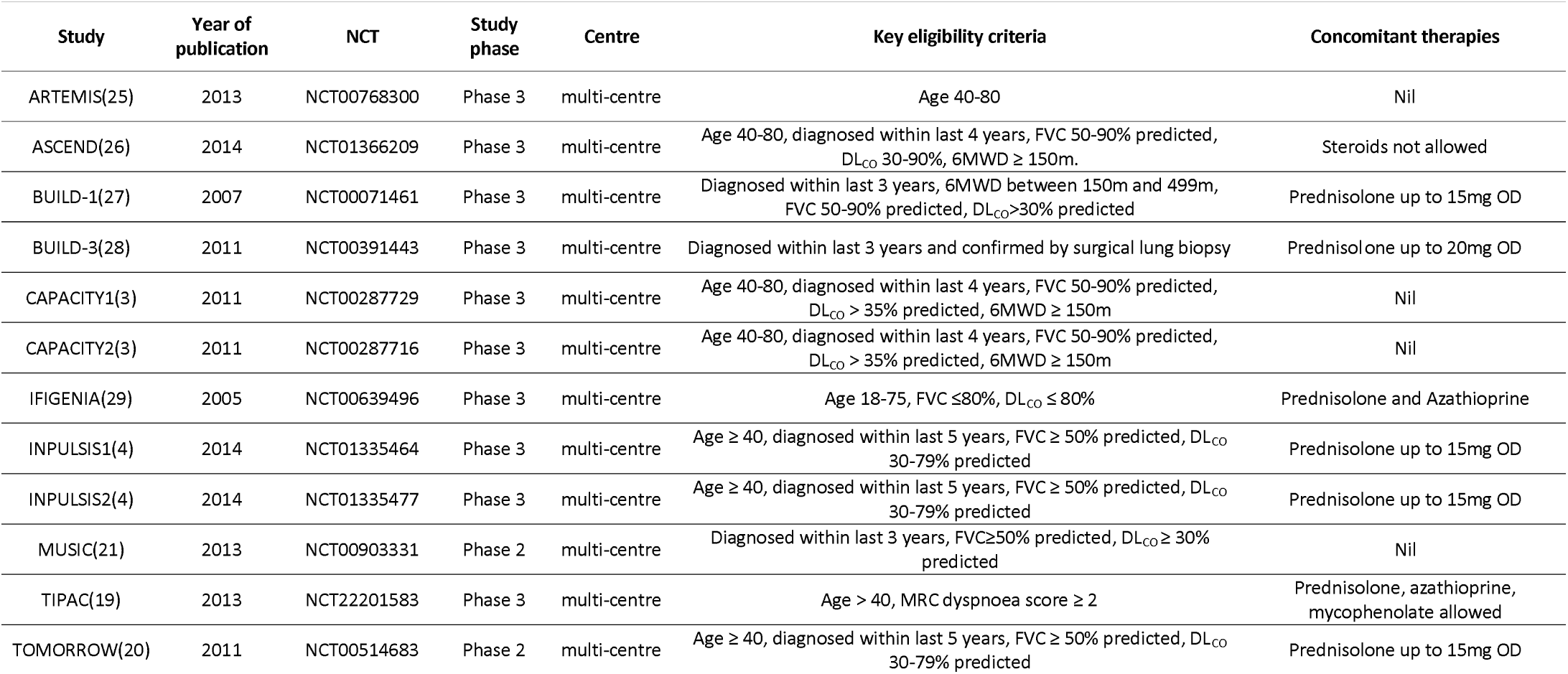
Methodological characteristics of included studies. 6MWD, six-minute walk distance, DL_CO_, gas transfer for carbon monoxide; FVC, forced vital capacity; MRC, medical research council

**Figure 1.**
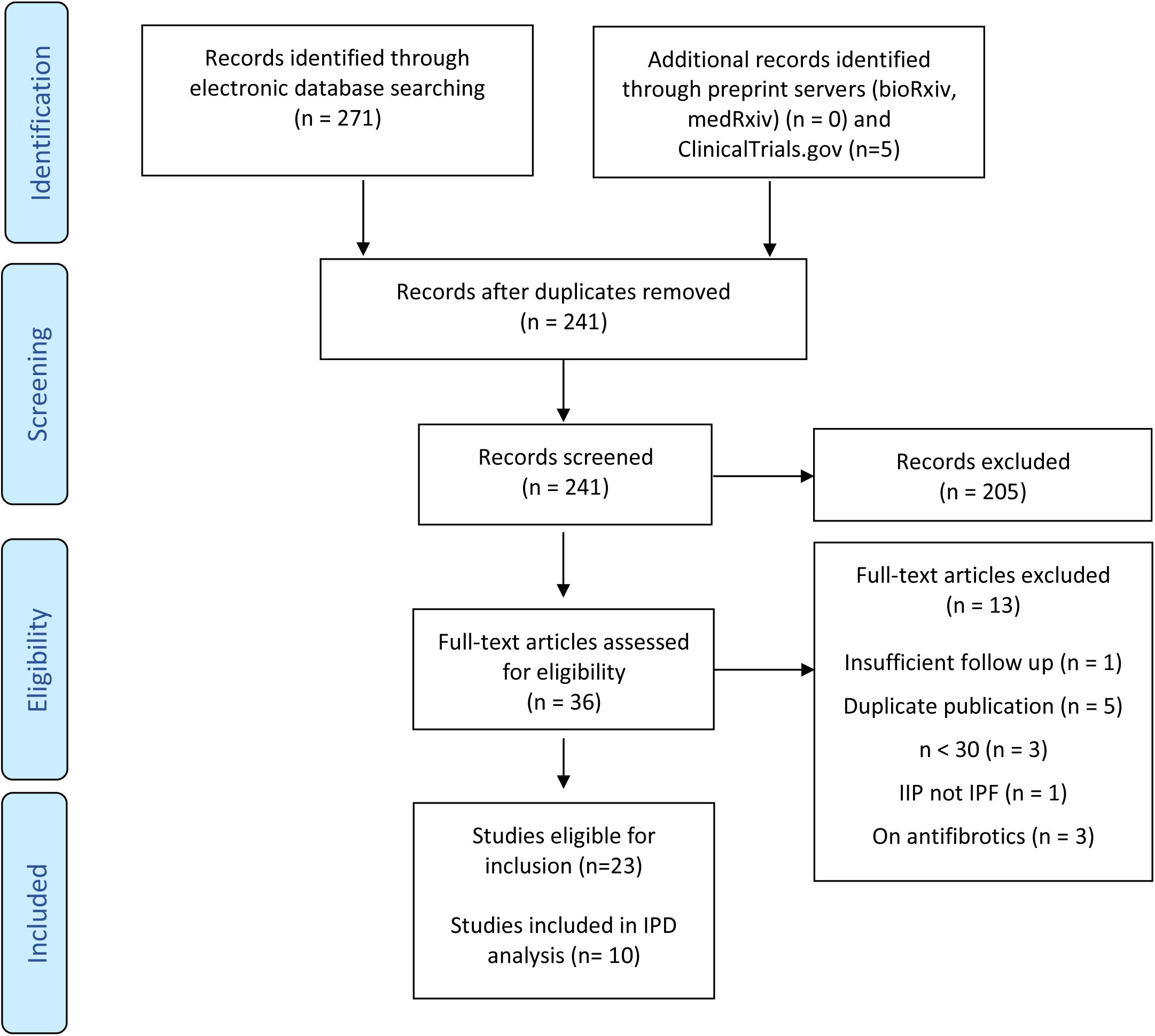
Search flow diagram Flow diagram illustrating systematic search and screening strategy, including numbers of studies meeting eligibility criteria and numbers excluded.

The median number of participants in the placebo arm of each study was 169 (IQR 85-207), with a median study age of 66.6 years (IQR 64.4-67.8) and median follow up duration of 13 months (IQR 12-17). Males represented 73.9% of all included participants. Associations of demographics with outcomes are provided in Figure E2. The median baseline %predicted FVC was 73.5% predicted (IQR 70.8-74.8), baseline %predicted DL_CO_ was 46.4% predicted (IQR 44.2-47.4), and 6MWD was 410m (IQR 365-421). Detailed study characteristics are provided in Table 2.

**Table 2.** Baseline participant characteristics. Baseline FVC % predicted values calculated using standardised global lung initiative (GLI) equations unless marked by asterisk (*). Values for physiological variables reported in mean (standard deviation) unless otherwise stated. 6MWD, six-minute walk distance, DL_CO_, gas transfer for carbon monoxide; FVC, forced vital capacity; MRC, medical research council; -, data not available

| Study | Placebo sample size | Study follow up, months (median, IQR) | Former or current smoker % | White ethnicity % | Age (years) | Sex – male (%) | Baseline FVC, L | Baseline FVC % predicted | Baseline DL<sub>CO</sub> % predicted | Baseline 6MWD |
| --- | --- | --- | --- | --- | --- | --- | --- | --- | --- | --- |
| ARTEMIS | 164 | 8 (5-13) | 67 | 89 | 66.1 (7.1) | 68.1 | - | 69.9 (13.8) * | 45.6 (13.3) | 421 (121) |
| ASCEND | 277 | 12 (11-12) | 61 | 90.6 | 67.8 (7.3) | 76.9 | 2.67 (0.65) | 70.8 (11.2) | 44.2 (12.5) | 421 (98) |
| BUILD-1 | 83 | 12 (12-12) | - | - | 70.2 (9.2) | 75 | 2.72 (0.72) | 73.9 (13.6) | 49.7 (11.3) | 365 (79) |
| BUILD-3 | 209 | 21 (15-24) | 67.9 | - | 63.2 (9.1) | 63.6 | 2.88 (0.82) | 73.1 (15.3) * | 47.9 (12.7) | - |
| CAPACITY1 | 174 | 17 (17-20) | 70.7 | 96.6 | 66.3 (7.5) | 73.6 | 2.91 (0.78) | 78 (15.5) | 46.1 (10.2) | 410 (90) |
| CAPACITY2 | 173 | 18 (17-22) | 63 | 98.8 | 67 (7.8) | 71.7 | 2.86 (0.68) | 75.3 (14.2) | 47.4 (9.2) | 400 (90) |
| IFIGENIA | 75 | 12 (11-13) | 69 | 100 | 64.4 (8.6) | 74.7 | 2.36 (0.73) | 62.8 (14.2) | 44.2 (15.9) | - |
| INPULSIS1 | 205 | 13 (13-13) | 75.1 | 80 | 66.8 (8.2) | 80 | 2.84 (0.82) | 75 (16.2) | 47.3 (11.9) | - |
| INPULSIS2 | 221 | 13 (13-13) | 67.4 | 56.2 | 67 (7.5) | 78.3 | 2.63 (0.8) | 72.8 (17) | 46.6 (15.4) | - |
| MUSIC | 65 | 13 (11-17) | 60 | 96.9 | 63.6 (6.1) | 61.5 | 2.79 (0.82) | 74.8 (14.6) * | 45.6 (11.2) | - |
| TIPAC | 86 | 12 (12-12) | 76.7 | 98.8 | 70.7 (8.6) | 65 | 2.4 (0.75) | 71.5 (21) | 39.1 (12.8) | 331 (118) |
| TOMORROW | 87 | 19 (14-23) | 66.7 | 77 | 64.8 (8.5) | 73.6 | 2.76 (0.74) | 74.6 (15) | 48.1 (13.1) | 410 (115) |

Risk of bias assessment identified a low risk of bias for most of the assessed domains (Figure 2, Table E1). All studies identified study participants with clear and consistent criteria. Reasons for participant drop out were provided in all studies and study attrition rates were low, except for one study which was terminated early due to futility. Details of physiological markers including methods of measurements and adopted FVC reference values were not provided in most studies. Important covariates such as age, sex and smoking status were consistently measured, and there was minimal missing outcome data in IPD, with mortality and disease progression status confidently defined in 99.95% and 87.1% of included participants respectively. Publication bias was detected by asymmetry of funnel plots for baseline FVC and DL_CO_ where mortality was estimated, and for change in FVC over three months for disease progression outcomes (Figures E11-E13)

**Figure 2.**
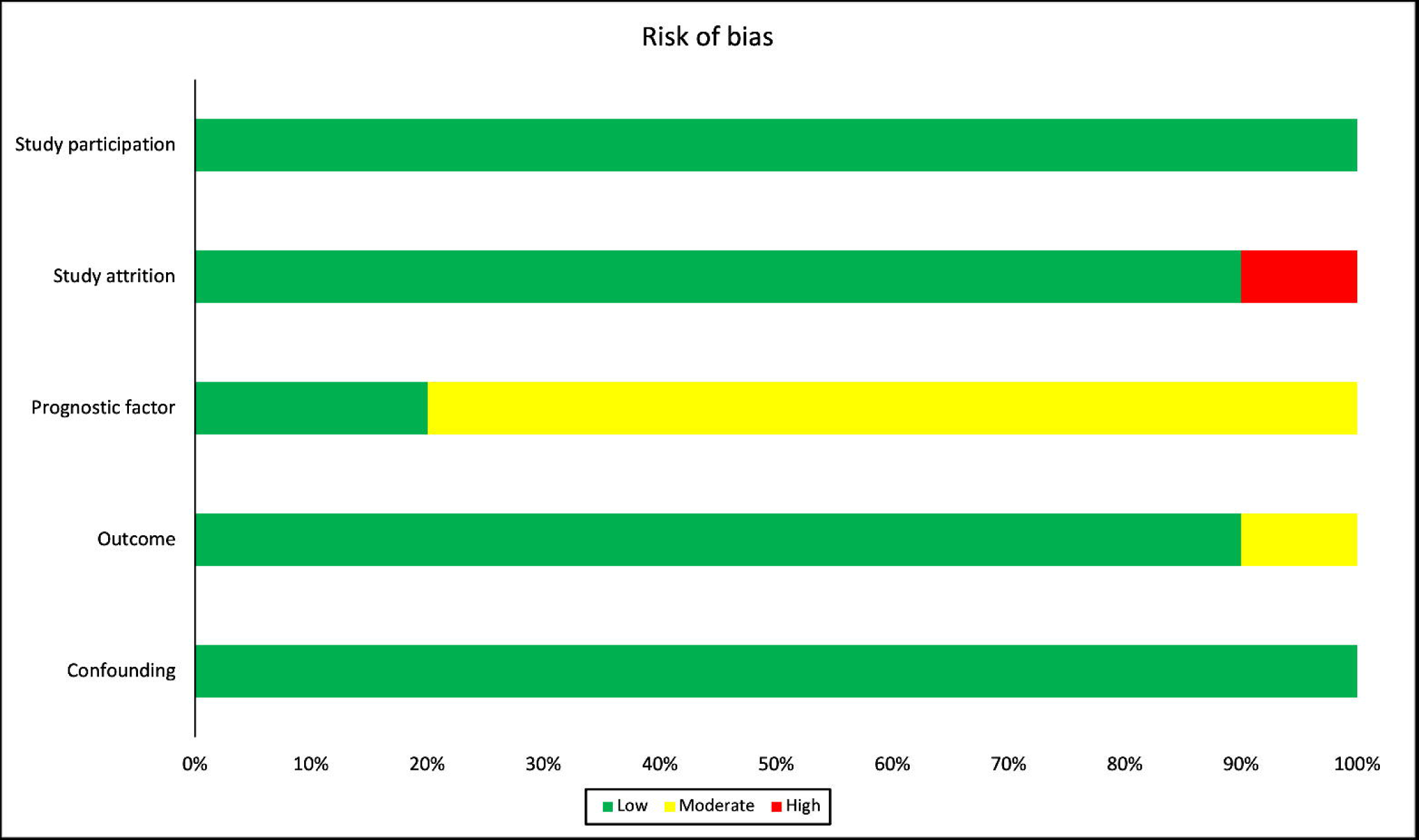
Risk of bias assessment. The risk of bias across studies was rated as low, moderate, or high risk in five categories using the modified QUIPs tool.

### Forced Vital Capacity (FVC)

FVC was available from all studies, with meta-analysis demonstrating, for every five point decrease in baseline %predicted FVC, there was an associated 24% increased risk of overall mortality [adjusted HR (aHR) 1.24 per FVC five point decrease in %predicted value, 95%CI 1.17;1.32, I^2^=0.0%, 11 cohorts, 1764 participants, moderate certainty] (Figure 3), and a 12% increased likelihood of disease progression [adjusted OR (aOR) 1.12 per FVC five point decrease in %predicted value, 95%CI 1.07;1.16, I^2^=0.0%, 11 cohorts, 1526 participants, high certainty]. The pooled absolute median decline in %predicted FVC from baseline to 12 months was 4.86% (95%CI -4.14;5.59, I^2^=68%). The mean FVC change at 12 months was - 201ml (95%CI -237; -164, I^2^=49.9%) in participants with a baseline FVC above 80% predicted, compared with -163ml (95%CI -201; -125, I^2^=74.4%) in individuals with a baseline FVC below 80% predicted (p=0.627), with a greater proportion of individuals dead at 12 months in those with a baseline FVC below 80% predicted (Figure E3).

**Figure 3.**
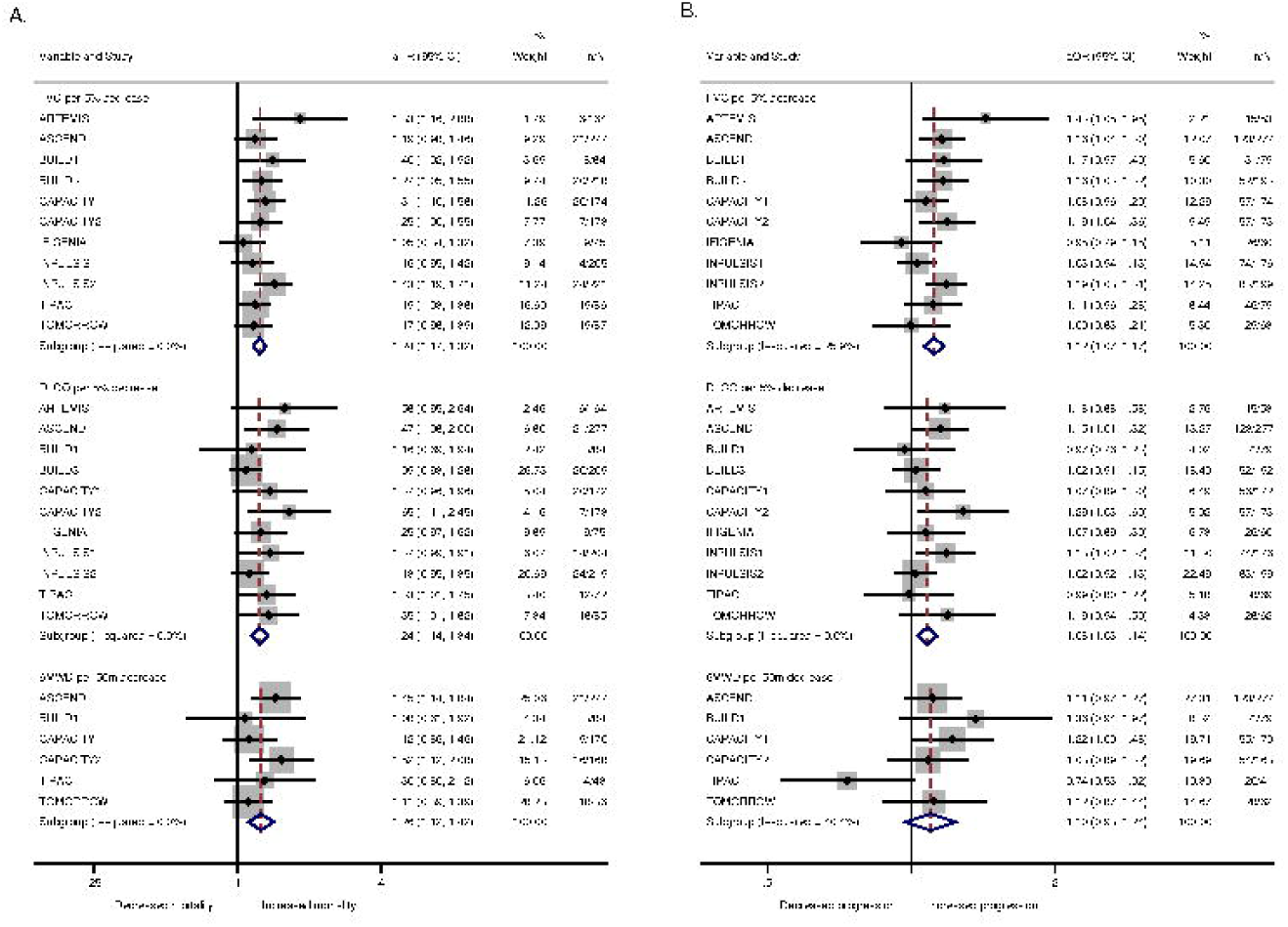
Forest plot of baseline physiological variables (continuous) and outcomes A: Adjusted hazard ratios (aHR) for overall mortality with 95% confidence intervals per 5% decline in lung function and 50m decline in 6MWD. Number of patients who died (n) alongside total patients included (N) in the study. All estimates were adjusted for age, sex and smoking status. B: Adjusted odds ratios (aOR) for disease progression with 95% confidence intervals per 5% decline in lung function and 50m decline in 6MWD. Number of progressors (n) alongside total patients included (N) in the study. All estimates were adjusted for age, sex and smoking status.

In analyses of three-month change, there was a 15% increased risk of overall mortality per 2.5% relative FVC decline (aHR 1.15 per 2.5% relative FVC decline in predicted value, 95%CI 1.06;1.24, I^2^ = 59.4%, 12 cohorts, 1729 participants, moderate certainty) (Figure 5), and 30% increased likelihood of disease progression (aOR 1.30 per 2.5% relative FVC decline in predicted value, 95%CI 1.19;1.41, I^2^=66.1%, 12 cohorts, 1551 participants, moderate certainty) (Figure 4). Meta-regression identified concomitant steroid use to be a source of heterogeneity for disease progression estimates in longitudinal FVC analysis (R^2^=31.65%; p=0.036) (Table E2). In subgroup analyses restricted to participants with a baseline FVC ≥ 80% predicted, three-month FVC change was associated with disease progression (aOR 1.48 per 2.5% relative FVC decline in predicted value, 95%CI 1.27;1.71, I^2^=0.0%, 412 participants), but not mortality (aHR 1.24 per 2.5% relative FVC decline in predicted value, 95%CI 0.92;1.67, I^2^=0.0%, 333 participants), though samples sizes were limited (Figure E4). The median three-month FVC decline in all included participants was 2.3%. A decline greater than 2.3% was associated with an estimated mean FVC difference of -280ml (95%CI -309; - 251, I^2^=43.7%) at 12months, compared with an estimated mean difference of -87ml (95%CI - 127; -48, I^2^=76.1%) in those participants with a lower three-month decline (p<0.001) (Figure E5).

**Figure 4.**
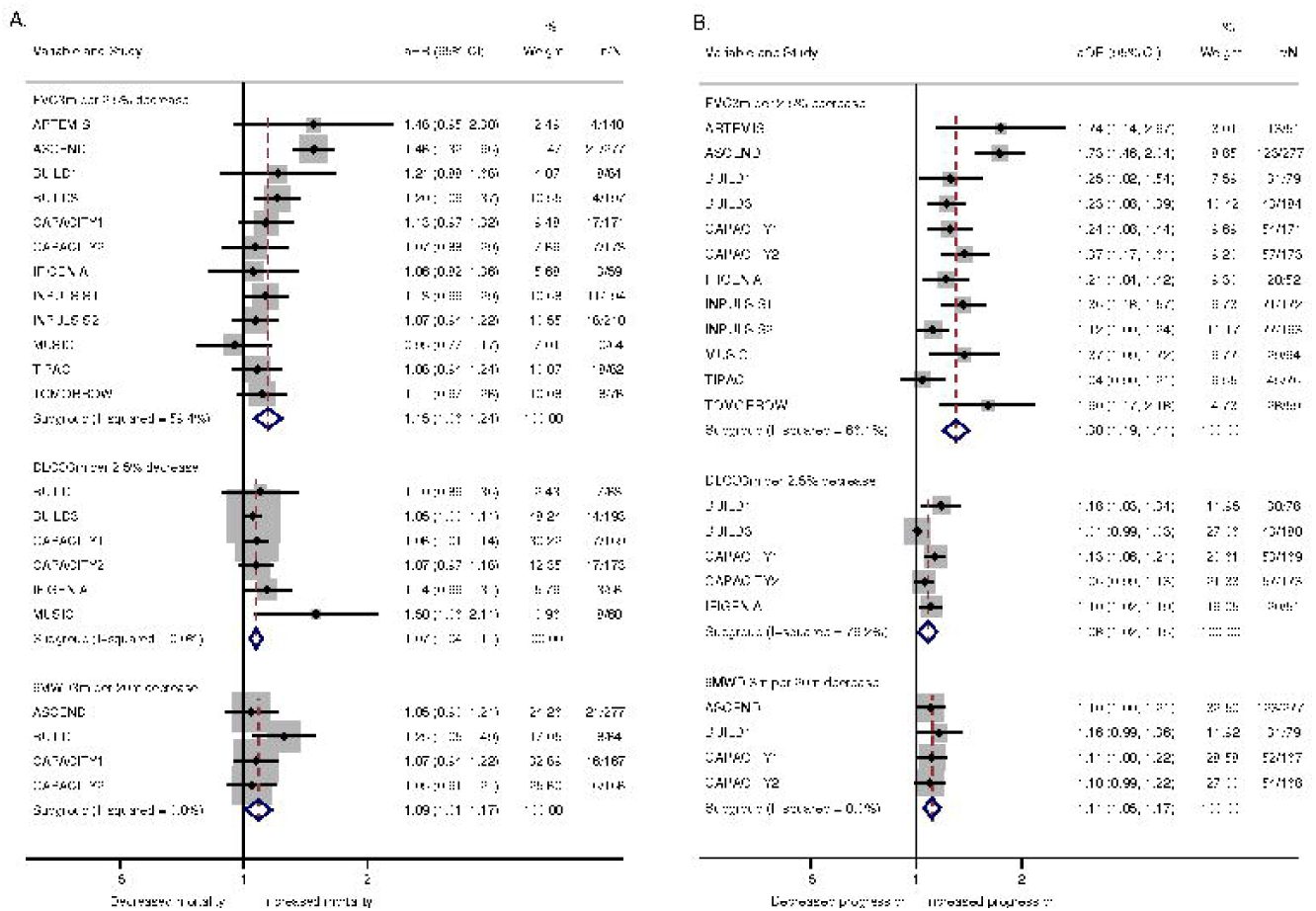
Forest plot of change in physiological variables (continuous) over three months and outcomes A: Adjusted hazard ratios (aHR) for overall mortality with 95% confidence intervals shown per 2.5% decline in lung function and 20m decline in 6MWD over 3 months. Number of patients who died (n) alongside total patients included (N) in the study. All estimates were adjusted for baseline values, age, sex and smoking status. B: Adjusted odds ratios (aOR) for disease progression with 95% confidence intervals shown per 2.5% decline in lung function and 20m decline in 6MWD over 3 months. Number of progressors (n) alongside total patients included (N) in the study. All estimates were adjusted for baseline values, age, sex and smoking status.

**Figure 5.**
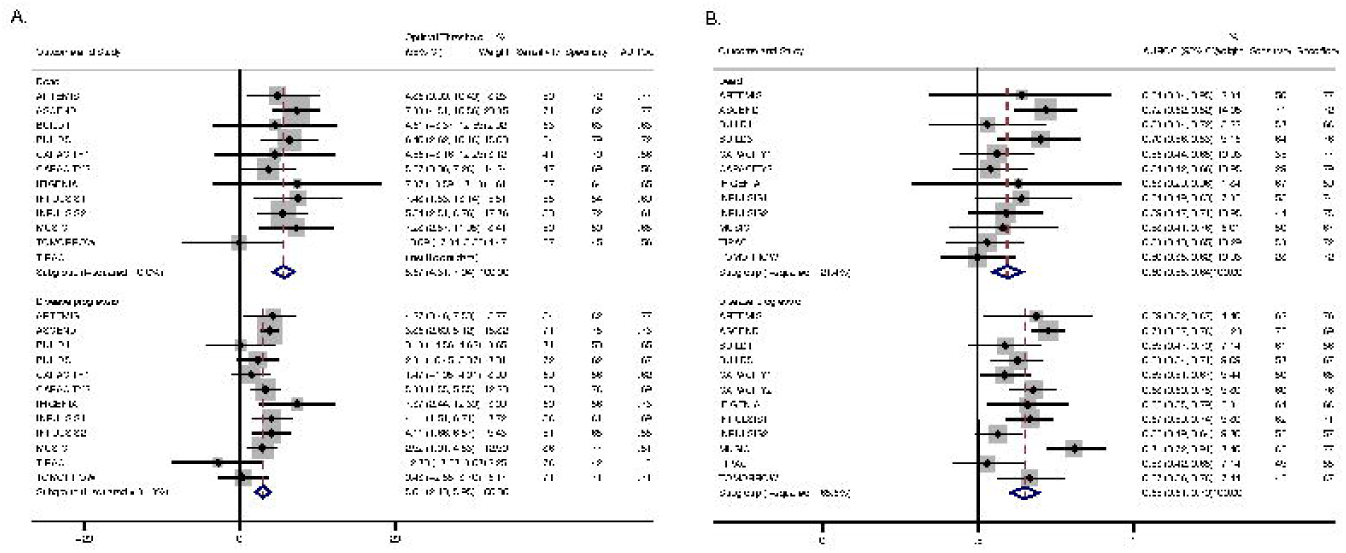
**A:** Forest plot of pooled optimal thresholds and 95% confidence intervals for 3-month relative FVC decline in predicting dead or disease progression. Optimal thresholds for 3-month relative FVC change in predicting outcomes were calculated and pooled to create an overall optimal threshold. **B:** Forest plot of AUROC for overall optimal threshold (5.7% for mortality and 3% for disease progression). The overall optimal threshold was applied to each study to calculate the AUROC, sensitivity and specificity for predicting outcomes. AUROC, area under receiver operating characteristics curve; Sensitivity and specificity in %.

Thresholds for determining the optimal three-month FVC relative change for distinguishing death and disease progression were calculated for each study independently and included in IPD meta-analysis (Figure 5). A pooled optimal threshold of 5.7% (95%CI 4.31;7.04, I^2^=0.0%, high certainty) was estimated for mortality, with an FVC decline greater than 5.7% over three months associated with a significantly increased risk of overall mortality compared with those who did not reach the threshold (aHR 2.62, 95%CI 1.73;3.96, I^2^=25.2%, 12 cohorts, 1734 participants, high certainty) (Figure E6). The pooled AUROC for this threshold was 0.60 (95%CI 0.55;0.64, I^2^=21.4%) (Figure 5). The optimal threshold for predicting disease progression was 3% (95%CI 2.10;3.93, I^2^=31.3%, high certainty), with an FVC decline greater than the threshold associated with an increased likelihood of disease progression compared with those who did not reach the threshold (OR 3.64, 95%CI 2.47;5.39, I^2^=58.5%, 12 cohorts, 1699 participants, moderate certainty) (Figure E7). Heterogeneity could not be explained by variability in any of the factors assessed. The pooled AUROC for predicting disease progression was 0.65 (95%CI 0.61;0.70, I^2^=65.6%), with concomitant steroid use associated with heterogeneity between studies (R^2^=34.74; p=0.041).

### Gas transfer for carbon monoxide (DL_CO_)

Baseline DL_CO_ was available across eleven cohorts (11/12), with availability of three-month DL_CO_ change limited to six cohorts (6/12). Meta-analysis estimated for every 5% decrease in baseline %predicted DLCO, there was an associated 24% increased risk of mortality (aHR 1.24, 95%CI 1.14;1.34, I^2^=0.0%, 11 cohorts, 1734 participants, moderate certainty) and an 8% increased likelihood of disease progression (aOR 1.08, 95%CI 1.03;1.14, I^2^=1.7%, 11 cohorts, 1512 participants, high certainty). In analyses over three months there was a 7% increased risk of mortality (aHR 1.07, 95%CI 1.04;1.11, I^2^=0.0%, 6 cohorts, 736 participants, moderate certainty) and 7% increase likelihood of disease progression (aOR 1.07; 95%CI 1.02;1.12, I^2^=70.8%, five cohorts, 651 participants, moderate certainty), per 2.5% DL_CO_ relative decline. Substantial heterogeneity was noted in the estimate of disease progression which could not be explained by variability in the factors assessed (Table E2). The optimal threshold for three-month relative decline in DL_CO_ was 10.51% (95%CI 4.14;16.88, I^2^=19.9%, 6 cohorts, low certainty) for predicting mortality (Figure E8 and E9), with an overall estimated AUROC of 0.64 (95%CI 0.54;0.74, I^2^=70.7%). The optimal threshold for predicting disease progression was 7.24% (95%CI 4.63;9.84, I^2^=0.0%, 5 cohorts, low certainty) with an overall estimated AUROC of 0.61(95%CI 0.57;0.66, I^2^=23.2%) (Figure E10).

### Six-minute walk distance (6MWD)

Baseline and three-month change in 6MWD were available from six (6/12) and four (4/12) cohorts respectively. Summary estimates from meta-analysis demonstrated there was a 26% increased risk of mortality (aHR 1.26, 95%CI 1.12;1.42, I^2^=0.0%, 6 cohorts, 828 participants, high certainty) and an inconclusive effect with disease progression (aOR 1.10, 95%CI 0.98;2.24, I^2^=40.4%, 5 cohorts, 718 participants, low certainty) per 50metre decrease in baseline walk distance. Although heterogeneity was low, meta-regression identified concomitant steroid use (R^2^=100%; p=0.006) to be a source of heterogeneity in mortality estimates, and the inclusion of participants with severe (R^2^=65.17; p=0.017) and non-incident cases (R^2^=65.17; p=0.017) as sources of heterogeneity in disease progression estimates. In longitudinal analyses, decline in 6MWD predicted mortality (aHR 1.09 per 20m decline, 95%CI 1.01;1.17, I^2^=0.0%, 4 cohorts, 696 participants, high certainty) and disease progression (aOR 1.11 per 20m decline; 95%CI 1.05;1.17, I^2^=0.0%, 4 cohorts, 691 participants, moderate certainty). Insufficient data precluded estimation of optimal three-month thresholds for predicting mortality and disease progression.

## DISCUSSION

This systematic review and IPD meta-analysis of placebo arms from interventional trials utilises robust methodology in well-characterised patients with untreated IPF to explore the association between commonly measured physiological variables and outcomes including mortality and disease progression. We report an independent association between three-month change in all physiological variables (FVC, DL_CO_, 6MWD), particularly FVC and poorer outcomes. A 15% increased risk of mortality and 30% increased likelihood of disease progression was estimated per 2.5% relative decline in FVC over three months. We used ROC analysis to identify optimal thresholds for three-month change in FVC that had the greatest sensitivity and specificity for predicting outcomes. All physiological variables measured at baseline were associated with mortality. We also evaluated demographic characteristics and found age, but not sex or previous smoking history, was associated with increased mortality. We assessed the certainty of our findings using GRADE (Table E3) and rated outcomes for both baseline and change in lung function with either moderate or high certainty depending on the outcome assessed.

Our findings have potential implications for the design of future clinical trials in IPF. Several included studies specified a primary endpoint as absolute change in FVC over 12 months. (4, 19-21) For example an FVC decline of 100ml (SD 300ml) would require a sample size of 310 participants using two sided tests to detect this endpoint (Cohen’s d 0.33) assuming 90% power, alpha 0.05, and equal allocation. A difference between arms of 5% relative decline over three months in %predicted FVC with a standard deviation of 15% in each arm offers the same effect size (Cohen’s d 0.33) to provide an additional early endpoint within the same sample size. Future studies could include this approach within study design, supporting refinement of the threshold and expected variability, as well as supporting objective evaluations in early study termination. An earlier endpoint would additionally support an adaptive design where experimental treatments can be added or dropped and multiple trials (phase 2 and 3) can be combined, increasing the efficiency of research in IPF. Sample size calculations for rate of change are particularly relevant in the anti-fibrotic era whereby cohort enrichment strategies are likely to be necessary due to slowing of disease progression and reduced event rates. The refinement of inclusion criteria to enrich for participants with a greater likelihood of progressive disease based on prior three-month FVC change may help further streamline interventional clinical trials.

Our findings support evidence that early changes in physiology may be surrogates for longer term prognosis, including two independent studies that identified an independent association between 24-week change in FVC and poorer outcomes in IPF(22, 23). Our study is the largest to evaluate the prognostic association of longitudinal change in physiological variables, and the first to establish the prognostic significance of change over a shorter time-period of 12-weeks, whilst concurrently identifying optimal threshold values.

Additionally, unlike previous pooled studies where participants from different studies were treated as one large cohort, this study is the first to utilise IPD meta-analysis to combine cohorts using a random effects model, and thus account for differences in individual trial populations.

The findings of this IPD meta-analysis should be interpreted in the context of several limitations. Our cohorts of patients were recruited in interventional clinical trials which carry a selection bias due to the exclusion of participants with severe disease or significant comorbidity at baseline, which can limit generalisability. Nonetheless it can be argued that change in physiological measurements add less value in advanced disease, and their principal benefit is the prognostication of non-severe disease. A further limitation was the requirement for participants to survive at least 12-weeks to be included in analyses of longitudinal physiology change, which may have underestimated our summary estimates. It is important to highlight existing trials with an endpoint of FVC change at 12 months face similar limitations with survival bias, often requiring imputation of missing data.(24) We were unable to consider an imputation algorithm to estimate missing values, as data from separate studies were accessed via multiple servers. A two-step IPD meta-analysis design was used to overcome the limitation of servers upon estimates. A further limitation was the dependence of secondary endpoints (disease progression and change in FVC at 12 months) on FVC as an exposure variable at earlier time points. However, findings for mortality, which was our primary endpoint, and for DL_CO_ and 6MWD were consistent. Three-month DL_CO_ was missing in 5/11 datasets, which could be non-random and due to severe disease, potentially underestimating effect estimates. IPD could not be retrieved from 1214 participants in 13/23 studies, raising the possibility of availability bias, although study and participants characteristics suggested little difference with included studies. Moreover, whether IPD was available from a particular trial was unlikely to be influenced by its findings, as the assessment of physiological variables as prognostic markers in placebo arms was not the objective of any trial. Searches were restricted to the English language introducing the possibility of missing eligible studies, although the possibility of this is small. We excluded cohorts where anti-fibrotic use was permitted, and future studies should assess the value of short-term change in physiology as a prognostic marker in patients receiving anti-fibrotic therapy.

In conclusion we conducted an IPD meta-analysis of RCT placebo arms to explore the association between physiological variables and important clinical endpoints in the largest, well-characterised cohorts of patients with untreated IPF. We demonstrate baseline and three-month change in physiological variables, particularly FVC, predict mortality, disease progression and change in FVC at 12 months. Our findings have the potential to streamline future clinical trials by utilising FVC change over three months as a surrogate endpoint.

## Data Availability

Meta-analysis of individual participant data from previously published papers. Original data available through data providers.

## Acknowledgements

This publication is based on research using data from data contributors Boehringer Ingelheim and Roche/Genentech Inc. that has been made available through Vivli, Inc. Vivli has not contributed to or approved, and is not in any way responsible for, the contents of this publication. Gilead Sciences, Inc. also contributed some data to this publication. Boehringer Ingelheim, Roche/Genentech, and Gilead Sciences Inc. were given the opportunity to review the abstract for medical and scientific accuracy, as well as intellectual property considerations. This publication also uses data from Actelion made available through Yale Open Data Access (YODA) Project, and data made available directly from ZAMBON Global Medical Affairs.

We would also like to thank Dr Allan Clark and Professor Andrew Wilson (University of East Anglia), for their invaluable support in providing access to individual patient data.

## Notes

Funding: FK is supported by the Nottingham National Institute for Health Research (NIHR) Biomedical Research Centre. RGJ is supported by an NIHR Research Professorship (RP-2017-08-ST2-014).

### Competing Interest Statement

The authors have declared no competing interest.

### Clinical Protocols

https://www.crd.york.ac.uk/prospero/display_record.php?RecordID=164935

### Funding Statement

FK is supported by the Nottingham National Institute for Health Research (NIHR) Biomedical Research Centre. RGJ is supported by an NIHR Research Professorship (RP-2017-08-ST2-014).

### Author Declarations

This meta-analysis makes use of individual participant data from studies where ethical approval and patient consent was obtained for sharing of data for these purposes. Furthermore all data were sought with signed data sharing agreements from data sharing partners which require strict concordance with rules pertaining to participant data. Ethical approval was not specifically sought for this meta-analysis.

## References

1. Navaratnam V, Fleming KM, West J, Smith CJ, Jenkins RG, Fogarty A, Hubbard RB. The rising incidence of idiopathic pulmonary fibrosis in the U.K. Thorax 2011; 66: 462–467.

2. Ley BC, H. R.// King, T. E., Jr. Clinical course and prediction of survival in idiopathic pulmonary fibrosis. Am J Respir Crit Care Med 2011; 183: 431–440.

3. Noble PW, Albera C, Bradford WZ, Costabel U, Glassberg MK, Kardatzke D, King TE, Jr., Lancaster L, Sahn SA, Szwarcberg J, Valeyre D, du Bois RM. Pirfenidone in patients with idiopathic pulmonary fibrosis (CAPACITY): two randomised trials. Lancet (London, England) 2011; 377: 1760–1769.

4. Richeldi L, du Bois RM, Raghu G, Azuma A, Brown KK, Costabel U, Cottin V, Flaherty KR, Hansell DM, Inoue Y, Kim DS, Kolb M, Nicholson AG, Noble PW, Selman M, Taniguchi H, Brun M, Le Maulf F, Girard M, Stowasser S, Schlenker-Herceg R, Disse B, Collard HR. Efficacy and safety of nintedanib in idiopathic pulmonary fibrosis. The New England journal of medicine 2014; 370: 2071–2082.

5. Stewart LA, Clarke M, Rovers M, Riley RD, Simmonds M, Stewart G, Tierney JF. Preferred Reporting Items for Systematic Review and Meta-Analyses of individual participant data: the PRISMA-IPD Statement. Jama 2015; 313: 1657–1665.

6. American Thoracic Society. Idiopathic pulmonary fibrosis: diagnosis and treatment. International consensus statement. American Thoracic Society (ATS), and the European Respiratory Society (ERS). Am J Respir Crit Care Med 2000; 161: 646–664.

7. American Thoracic Society/European Respiratory Society International Multidisciplinary Consensus Classification of the Idiopathic Interstitial Pneumonias. This joint statement of the American Thoracic Society (ATS), and the European Respiratory Society (ERS) was adopted by the ATS board of directors, June 2001 and by the ERS Executive Committee, June 2001. Am J Respir Crit Care Med 2002; 165: 277–304.

8. Raghu G, Collard HR, Egan JJ, Martinez FJ, Behr J, Brown KK, Colby TV, Cordier JF, Flaherty KR, Lasky JA, Lynch DA, Ryu JH, Swigris JJ, Wells AU, Ancochea J, Bouros D, Carvalho C, Costabel U, Ebina M, Hansell DM, Johkoh T, Kim DS, King TE, Jr., Kondoh Y, Myers J, Muller NL, Nicholson AG, Richeldi L, Selman M, Dudden RF, Griss BS, Protzko SL, Schunemann HJ, Fibrosis AEJACoIP. An official ATS/ERS/JRS/ALAT statement: idiopathic pulmonary fibrosis: evidence-based guidelines for diagnosis and management. Am J Respir Crit Care Med 2011; 183: 788–824.

9. www.vivli.org.

10. https://yoda.yale.edu. The Yoda Project.

11. www.clinicalstudydatarequest.com.

12. Hayden JA, van der Windt DA, Cartwright JL, Cote P, Bombardier C. Assessing bias in studies of prognostic factors. Ann Intern Med 2013; 158: 280–286.

13. Iorio A, Spencer FA, Falavigna M, Alba C, Lang E, Burnand B, McGinn T, Hayden J, Williams K, Shea B, Wolff R, Kujpers T, Perel P, Vandvik PO, Glasziou P, Schunemann H, Guyatt G. Use of GRADE for assessment of evidence about prognosis: rating confidence in estimates of event rates in broad categories of patients. BMJ : British Medical Journal 2015; 350: h870.

14. Burke DL, Ensor J, Riley RD. Meta-analysis using individual participant data: one-stage and two-stage approaches, and why they may differ. Stat Med 2017; 36: 855–875.

15. Cooper BG, Stocks J, Hall GL, Culver B, Steenbruggen I, Carter KW, Thompson BR, Graham BL, Miller MR, Ruppel G, Henderson J, Vaz Fragoso CA, Stanojevic S. The Global Lung Function Initiative (GLI) Network: bringing the world’s respiratory reference values together. Breathe (Sheff) 2017; 13: e56–e64.

16. NICE. Idiopathic pulmonary fibrosis in adults: diagnosis and management; 2013.

17. Liu X. Classification accuracy and cut pointllselection. Statistics in Medicine 2012; 31: 2676–2686.

18. Egger M, Smith GD, Schneider M, Minder C. Bias in meta-analysis detected by a simple, graphical test. BMJ 1997; 315: 629–634.

19. Shulgina L, Cahn AP, Chilvers ER, Parfrey H, Clark AB, Wilson ECF, Twentyman OP, Davison AG, Curtin JJ, Crawford MB, Wilson AM. Treating idiopathic pulmonary fibrosis with the addition of co-trimoxazole: a randomised controlled trial. Thorax 2013; 68: 155–162.

20. Richeldi L, Costabel U, Selman M, Kim DS, Hansell DM, Nicholson AG, Brown KK, Flaherty KR, Noble PW, Raghu G, Brun M, Gupta A, Juhel N, Klüglich M, du Bois RM. Efficacy of a tyrosine kinase inhibitor in idiopathic pulmonary fibrosis. The New England journal of medicine 2011; 365: 1079–1087.

21. Raghu G, Million-Rousseau R, Morganti A, Perchenet L, Behr J. Macitentan for the treatment of idiopathic pulmonary fibrosis: the randomised controlled MUSIC trial. The European respiratory journal 2013; 42: 1622–1632.

22. Ley B, Bradford WZ, Vittinghoff E, Weycker D, du Bois RM, Collard HR. Predictors of Mortality Poorly Predict Common Measures of Disease Progression in Idiopathic Pulmonary Fibrosis. Am J Respir Crit Care Med 2016; 194: 711–718.

23. du Bois RM, Weycker D, Albera C, Bradford WZ, Costabel U, Kartashov A, Lancaster L, Noble PW, Raghu G, Sahn SA, Szwarcberg J, Thomeer M, Valeyre D, King TE, Jr. Ascertainment of individual risk of mortality for patients with idiopathic pulmonary fibrosis. Am J Respir Crit Care Med 2011; 184: 459–466.

24. King TE, Jr., Bradford WZ, Castro-Bernardini S, Fagan EA, Glaspole I, Glassberg MK, Gorina E, Hopkins PM, Kardatzke D, Lancaster L, Lederer DJ, Nathan SD, Pereira CA, Sahn SA, Sussman R, Swigris JJ, Noble PW, Group AS. A phase 3 trial of pirfenidone in patients with idiopathic pulmonary fibrosis. New England Journal of Medicine 2014; 370: 2083–2092.

25. Raghu G, Behr J, Brown KK, Egan JJ, Kawut SM, Flaherty KR, Martinez FJ, Nathan SD, Wells AU, Collard HR, Costabel U, Richeldi L, de Andrade J, Khalil N, Morrison LD, Lederer DJ, Shao L, Li X, Pedersen PS, Montgomery AB, Chien JW, O’Riordan TG. Treatment of idiopathic pulmonary fibrosis with ambrisentan: a parallel, randomized trial. Ann Intern Med 2013; 158: 641–649.

26. King TE, Jr., Bradford WZ, Castro-Bernardini S, Fagan EA, Glaspole I, Glassberg MK, Gorina E, Hopkins PM, Kardatzke D, Lancaster L, Lederer DJ, Nathan SD, Pereira CA, Sahn SA, Sussman R, Swigris JJ, Noble PW. A phase 3 trial of pirfenidone in patients with idiopathic pulmonary fibrosis. The New England journal of medicine 2014; 370: 2083–2092.

27. King TE, Jr., Behr J, Brown KK, du Bois RM, Lancaster L, de Andrade JA, Stähler G, Leconte I, Roux S, Raghu G. BUILD-1: a randomized placebo-controlled trial of bosentan in idiopathic pulmonary fibrosis. Am J Respir Crit Care Med 2008; 177: 75–81.

28. King TE, Jr., Brown KK, Raghu G, du Bois RM, Lynch DA, Martinez F, Valeyre D, Leconte I, Morganti A, Roux S, Behr J. BUILD-3: a randomized, controlled trial of bosentan in idiopathic pulmonary fibrosis. Am J Respir Crit Care Med 2011; 184: 92–99.

29. Behr J, Demedts M, Buhl R, Costabel U, Dekhuijzen RP, Jansen HM, MacNee W, Thomeer M, Wallaert B, Laurent F, Nicholson AG, Verbeken EK, Verschakelen J, Flower CD, Petruzzelli S, De Vuyst P, van den Bosch JM, Rodriguez-Becerra E, Lankhorst I, Sardina M, Boissard G. Lung function in idiopathic pulmonary fibrosis--extended analyses of the IFIGENIA trial. Respir Res 2009; 10: 101.

