## Supplementary material for "Three month FVC change: a trial endpoint for IPF based on individual participant data meta-analysis"

### Online Data Supplement

Figure E1 – MEDLINE search strategy

Figure E2 - Forest plot of baseline demographic variables (continuous) and outcomes

Figure E3 – Forest plot for 12m FVC (ml) change stratified by baseline FVC

Figure E4 – Forest plot for 3-month FVC change in predicting outcomes in those with a baseline FVC ≥ 80%

Figure E5 - Forest plot for 12m FVC (ml) change stratified by 3m FVC change

Figure E6 – Forest plot for 3-month FVC empirical mortality threshold (5.7%) applied to all studies.

Figure E7 – Forest plot for 3-month FVC empirical disease progression threshold (3%) applied to all studies

Figure E8 – Forest plot for 3-month DLCO threshold change

Figure E9 – Forest plot for 3-month $DL_{CO}$ empirical mortality threshold

Figure E10 - Forest plot for 3-month $DL_{CO}$ empirical disease progression threshold

Figure E11 - FVC publication bias

Figure E12 – DLCO publication bias

Figure E13 – 6MWD publication bias

Table E1 – Risk of bias assessment

Table E2 – Meta-regression

Table E3 – GRADE assessment

Table E4 – Baseline participant characteristics from studies not included in meta-analysis

1. idiopathic pulmonary fibros*.mp.
2. pulmonary fibros*.mp.
3. Pulmonary Fibrosis/ or Idiopathic Pulmonary Fibrosis/
4. cryptogenic fibrosing alveolitis.mp.
5. usual interstitial pneumonia*.mp.
6. Fibrosing alveolitis.mp.
7. Idiopathic Interstitial Pneumonia*.mp.
8. Interstitial pneumonia*.mp.
9. Idiopathic interstitial lung disease.mp.
10. Chronic interstitial pneumonia*.mp.
11. 1 or 2 or 3 or 4 or 5 or 6 or 7 or 8 or 9 or 10
12. Forced Vital Capacity.mp. or Vital Capacity/
13. FVC.mp.
14. Forced Expiratory Volume/ or FEV1.mp.
15. forced expiratory volume.mp.
16. 6 minute walk.mp.
17. Six minute walk.mp.
18. Walk Test/
19. walk test.mp.
20. 6MWT.mp.
21. 6MWD.mp.
22. Pulmonary diffusing capacity.mp. or Pulmonary Diffusing Capacity/
23. Diffusion capacity for carbon monoxide.mp.
24. DLCO.mp.
25. Transfer factor.mp. or Transfer Factor/
26. Gas transfer.mp.
27. TLCO.mp.
28. KCO.mp.
29. PHYSIOLOGY/
30. Physiolog*.mp.
31. SPIROMETRY/
32. spiromet*.mp.
33. 12 or 13 or 14 or 15 or 16 or 17 or 18 or 19 or 20 or 21 or 22 or 23 or 24 or 25 or 26 or 27 or 28 or 29 or 30 or 31 or 32
34. 11 and 33
35. limit 34 to humans

36.. limit 35 to english language

37.. limit 36 to (clinical trial, all or randomized controlled trial)

**Figure E1** – MEDLINE search strategy (last search carried out 1st Dec 2020)

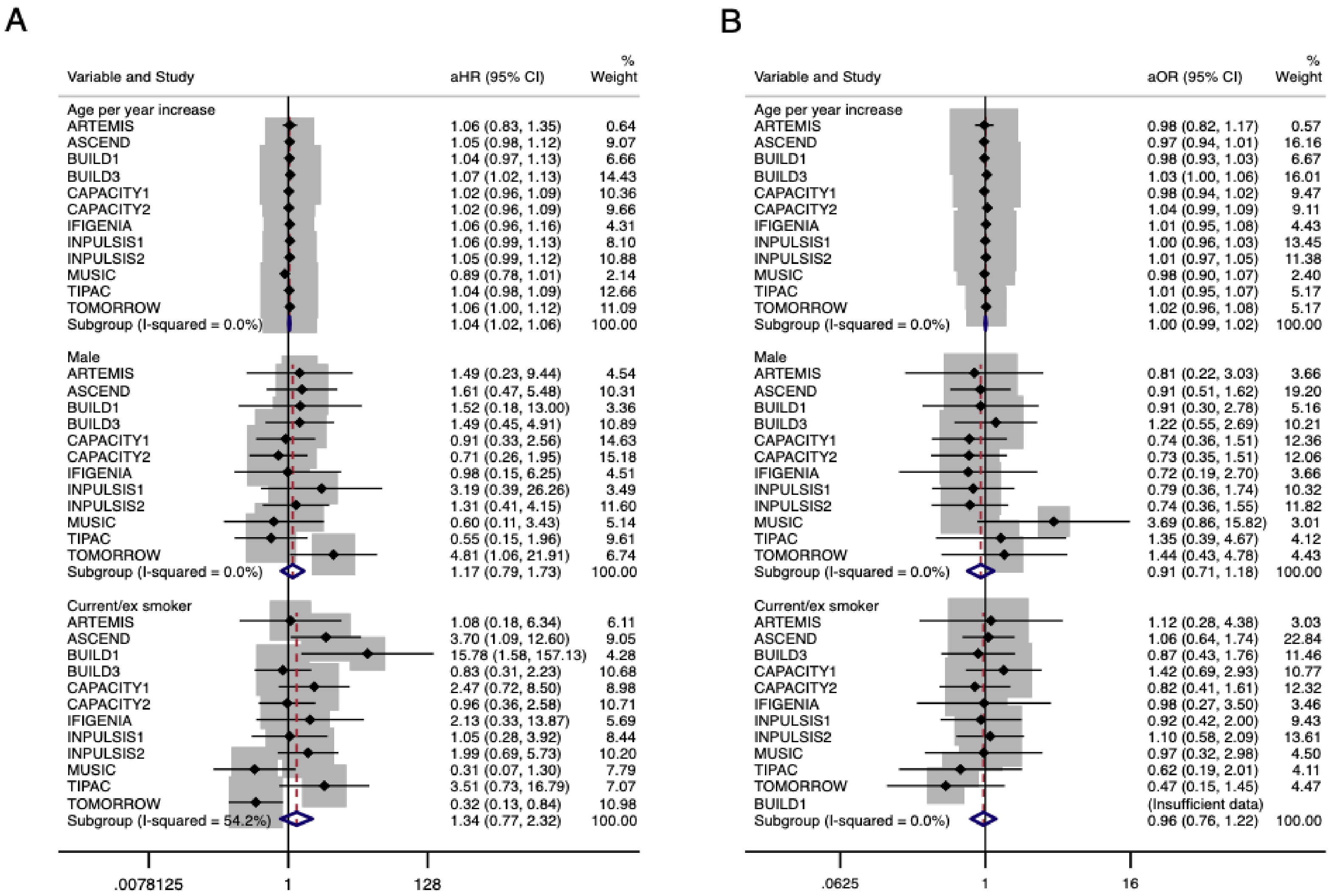


**Figure E2** – Forest plot of baseline demographic variables (continuous) and outcomes

A: Adjusted hazard ratios (aHR) for overall mortality with 95% confidence intervals shown for each study and pooled estimates

B: Adjusted odds ratios (aOR) for disease progression with 95% confidence intervals shown for each study and pooled estimates

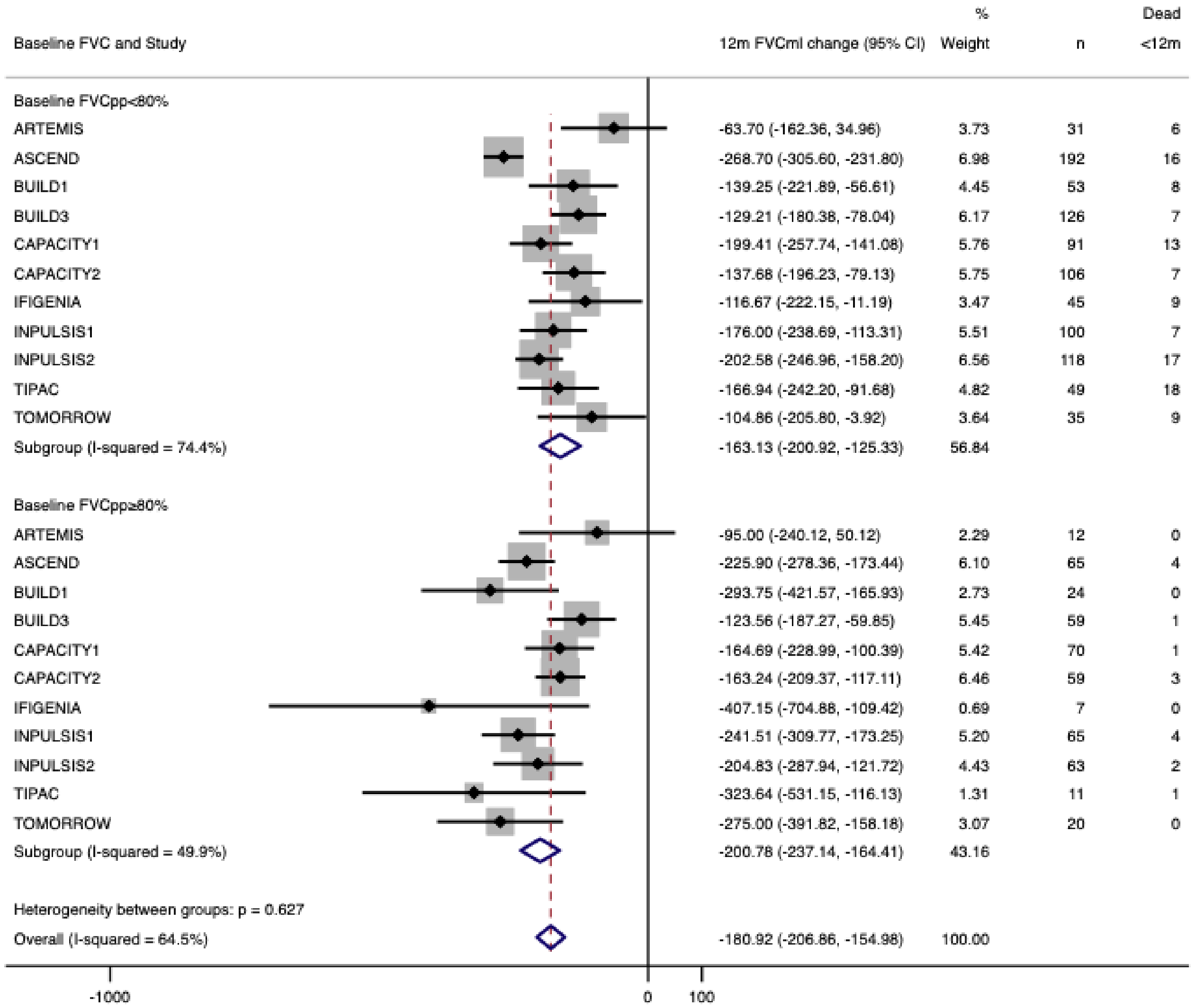


**Figure E3** – Forest plot for 12m FVC (ml) change stratified by baseline FVC. Baseline FVC was stratified by a threshold of 80% predicted and FVC (ml) change at 12m was calculated and compared in both groups.

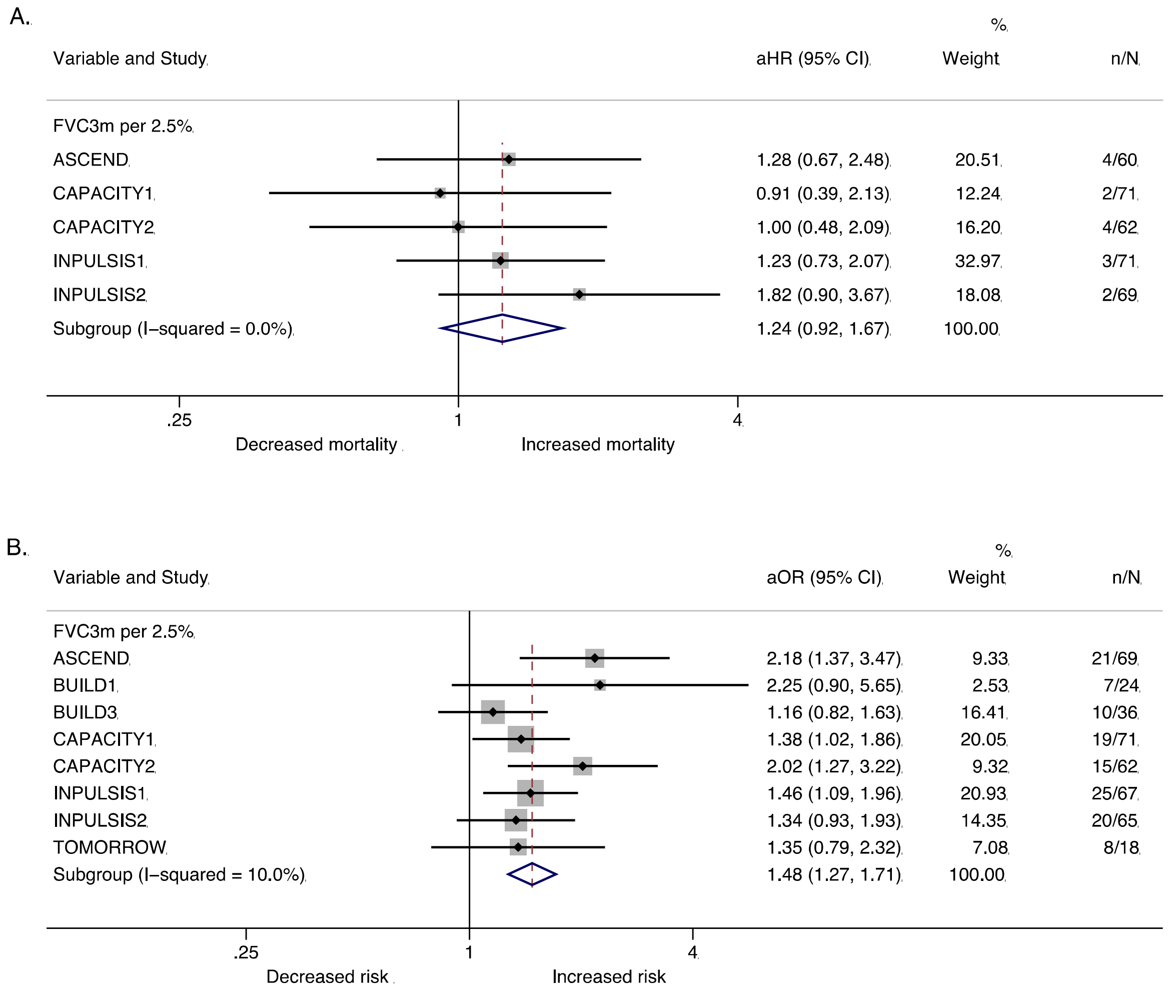


**Figure E4** – Forest plot for 3-month FVC change in participants with baseline FVC ≥ 80% predicted only. A. Adjusted hazard ratios (aHR) for overall mortality with 95% CI shown per 2.5% decline in FVC. Number of patients who died (n) alongside total patients included (N) in the analysis. B: Adjusted odds ratios (aOR) for disease progression with 95% CI shown per 2.5% decline in FVC over 3 months. Number of progressors (n) alongside total patients included (N) in the analysis. All estimates were adjusted for baseline FVC, age, sex, and smoking status.

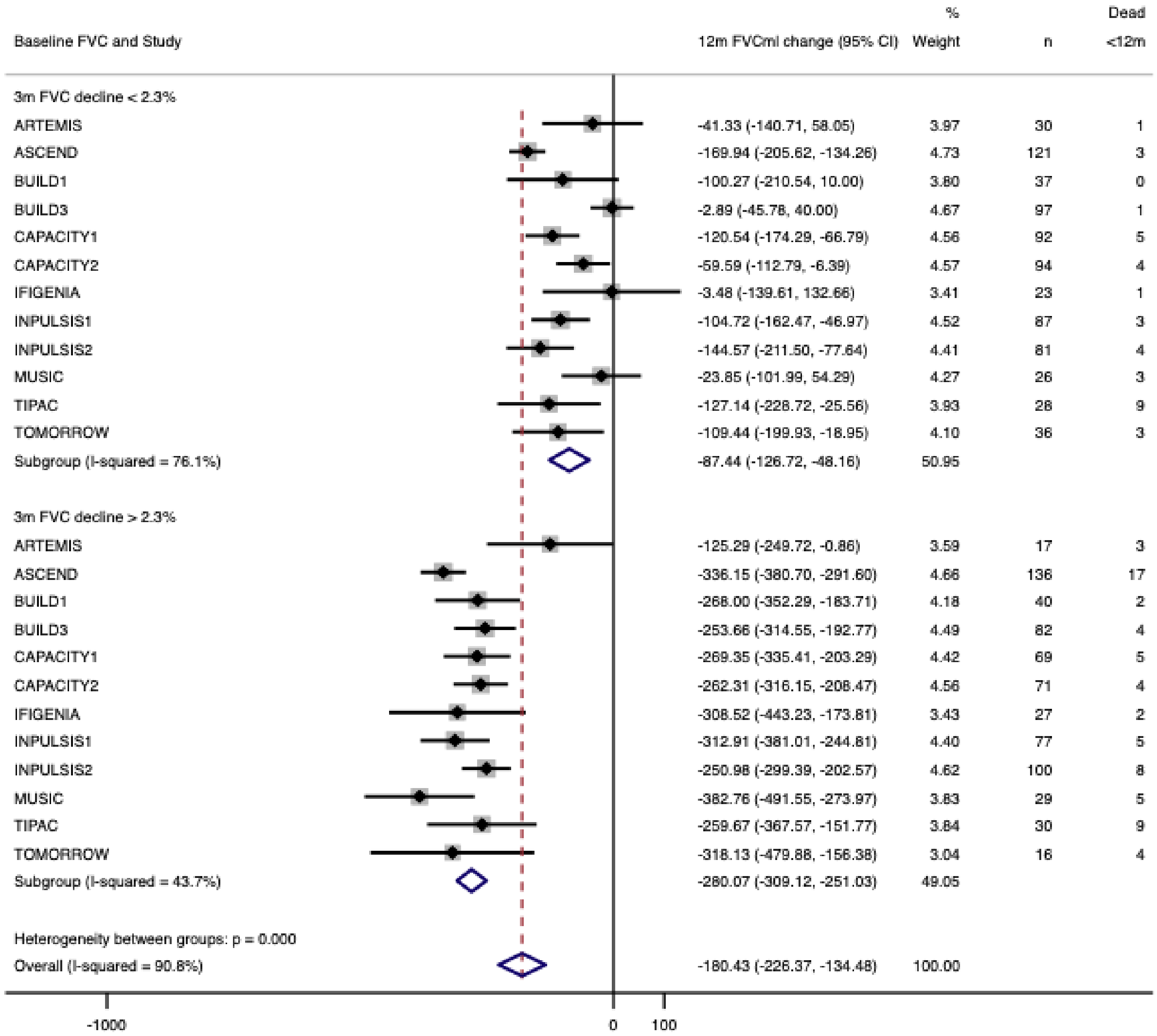


**Figure E5** - Forest plot for 12m FVC (ml) change stratified by 3m FVC change. The median FVC change for all participants was calculated (2.3%) and mean FVC (ml) change at 12m was calculated in participants with a greater than 3m threshold change compared with a change below the threshold.

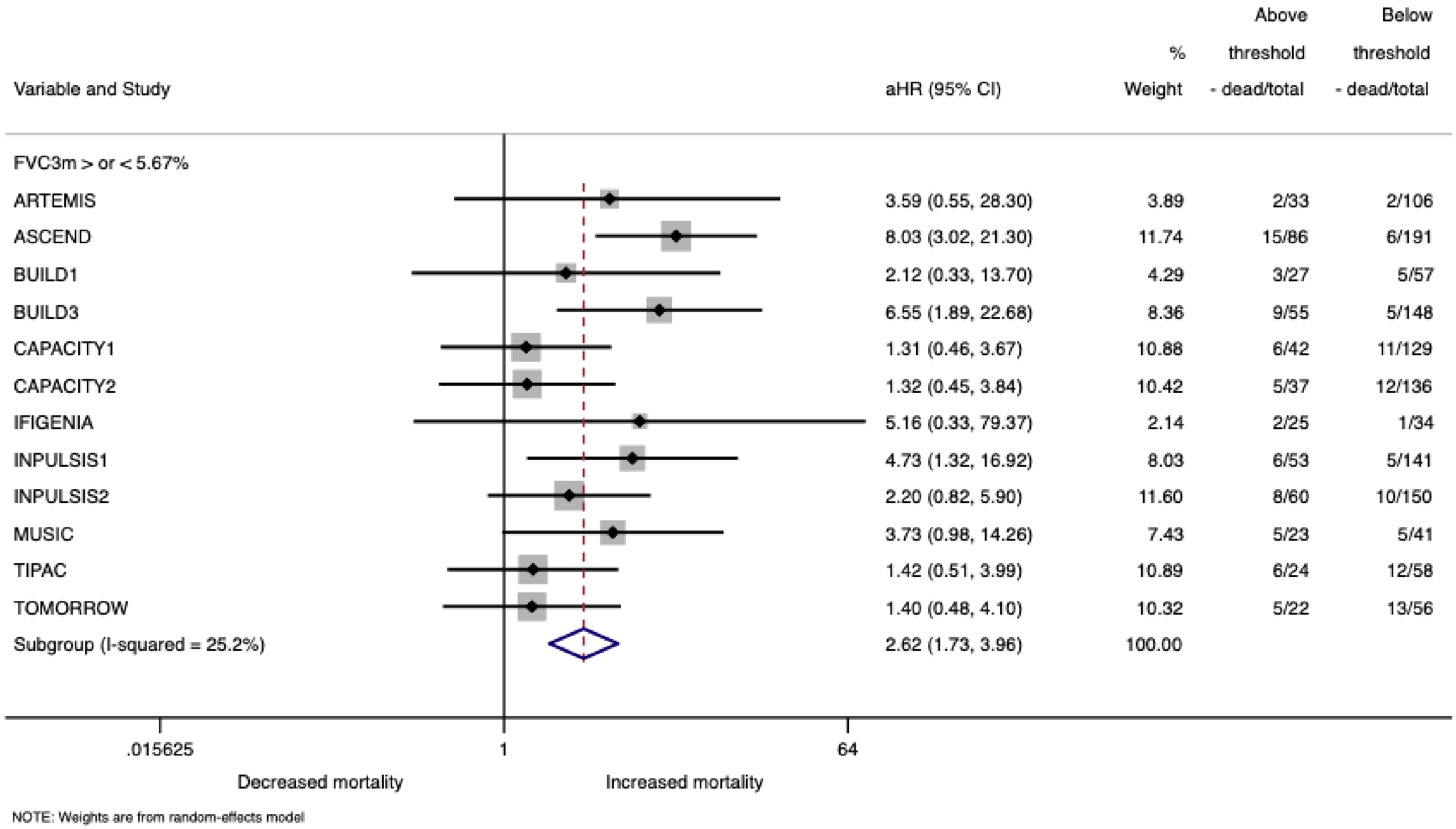


**Figure E6** – Forest plot for 3-month FVC empirical mortality threshold (5.7%) applied to all studies. An optimum threshold for 3-month relative FVC change in predicting mortality was calculated and pooled. The pooled threshold (5.7%) was applied to all studies to estimate the risk of overall mortality in patients with an FVC decline greater than 5.7% predicted over three months compared with patients who had an FVC decline less than 5.7% predicted.

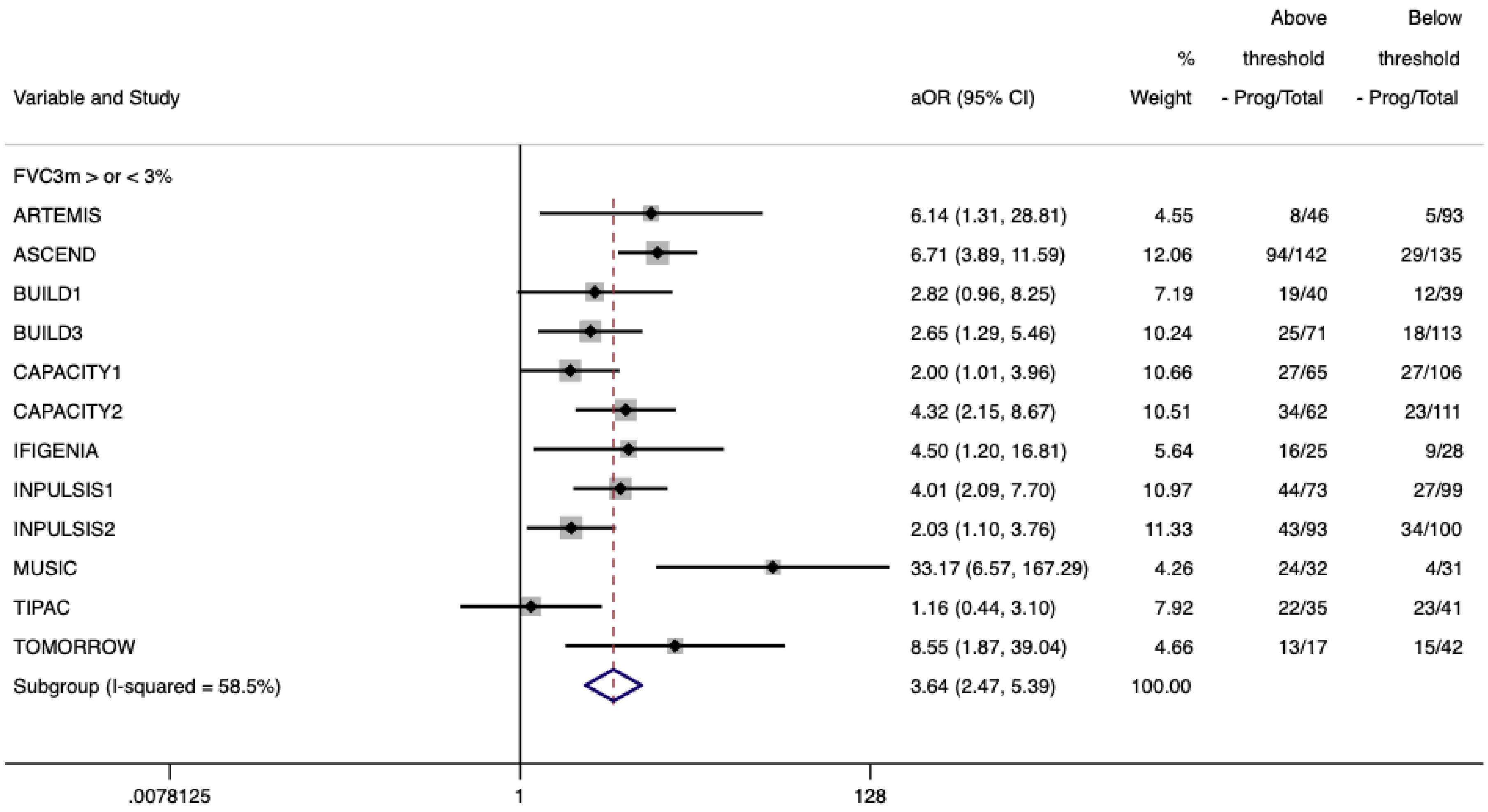


**Figure E7** – Forest plot for 3-month FVC empirical disease progression threshold (3%) applied to all studies. An optimum threshold for 3-month relative FVC change in predicting disease progression was calculated and pooled. The pooled threshold (3%) was applied to all studies to estimate the risk of overall disease progression in patients with an FVC decline greater than 3% predicted over three months compared with patients who had an FVC decline less than 3% predicted.

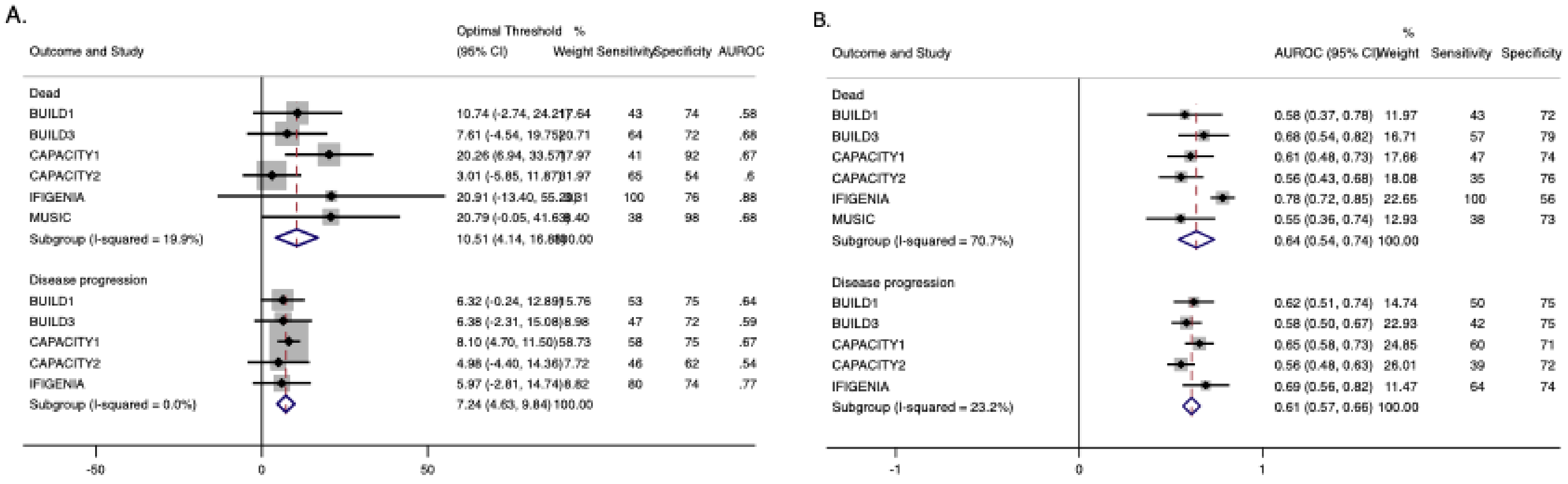


**Figure E8 - A:** Forest plot of pooled optimal thresholds and 95% confidence intervals for 3-month relative $DL_{CO}$ decline in predicting dead or disease progression. Optimal thresholds for 3-month relative $DL_{CO}$ change in predicting outcomes were calculated and pooled to create an overall optimal threshold.

**B:** Forest plot of AUROC for overall optimal threshold (10.5% for mortality and 7.2% for disease progression). The overall optimal threshold was applied to each study to calculate the AUROC, sensitivity and specificity for predicting outcomes.

AUROC, area under receiver operating characteristics curve; Sensitivity and specificity in %.

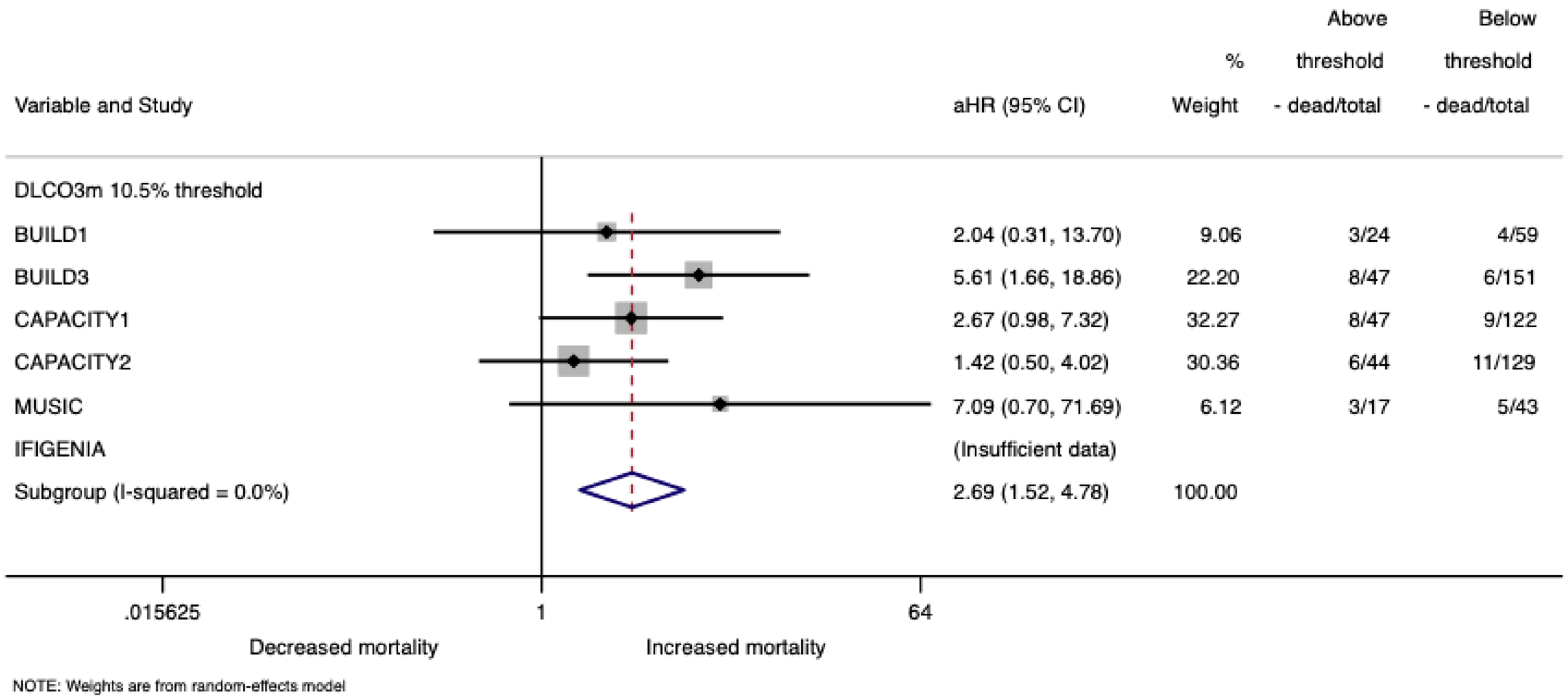


**Figure E9** – Forest plot for 3-month $DL_{CO}$ empirical mortality threshold (10.5%) applied to all studies. An optimum threshold for 3-month relative $DL_{CO}$ change in predicting mortality was calculated and pooled. The pooled threshold (10.5%) was applied to all studies to estimate the risk of overall mortality in patients with an $DL_{CO}$ decline greater than 10.5% predicted over three months compared with patients who had an $DL_{CO}$ decline less than 10.5% predicted.

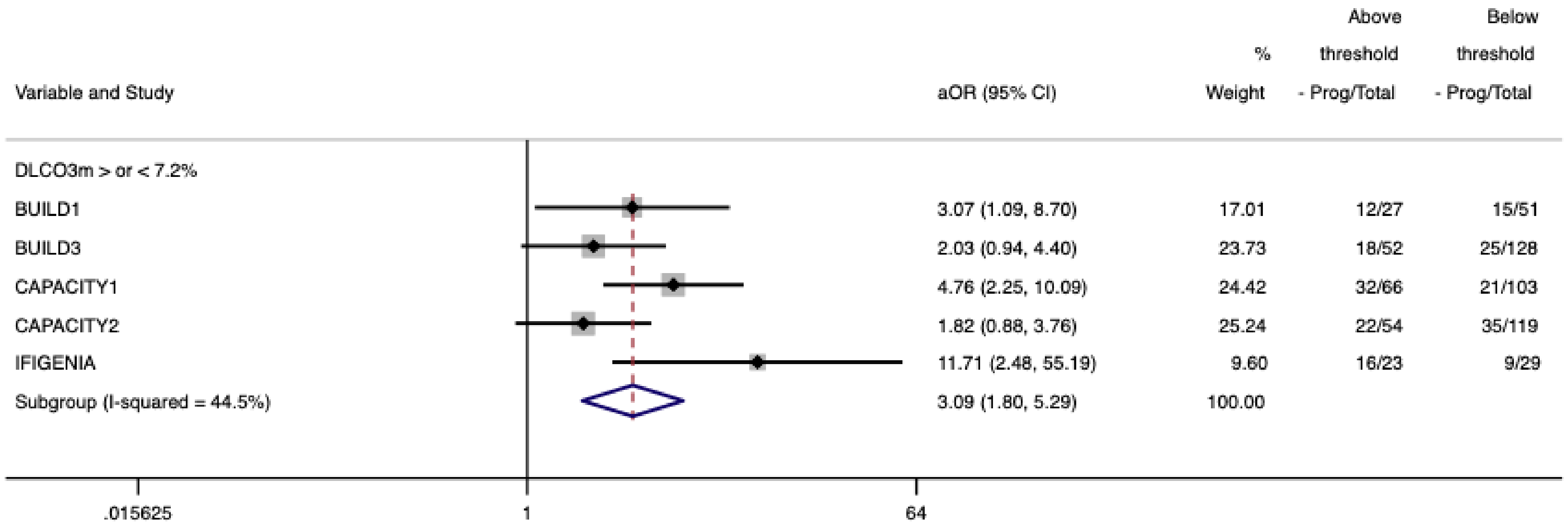


**Figure E10** – Forest plot for 3-month $DL_{CO}$ empirical disease progression threshold (7.2%) applied to all studies. An optimum threshold for 3-month relative $DL_{CO}$ change in predicting disease progression was calculated and pooled. The pooled threshold (7.2%) was applied to all studies to estimate the risk of overall disease progression in patients with an $DL_{CO}$ decline greater than 7.2% predicted over three months compared with patients who had an $DL_{CO}$ decline less than 7.2% predicted.

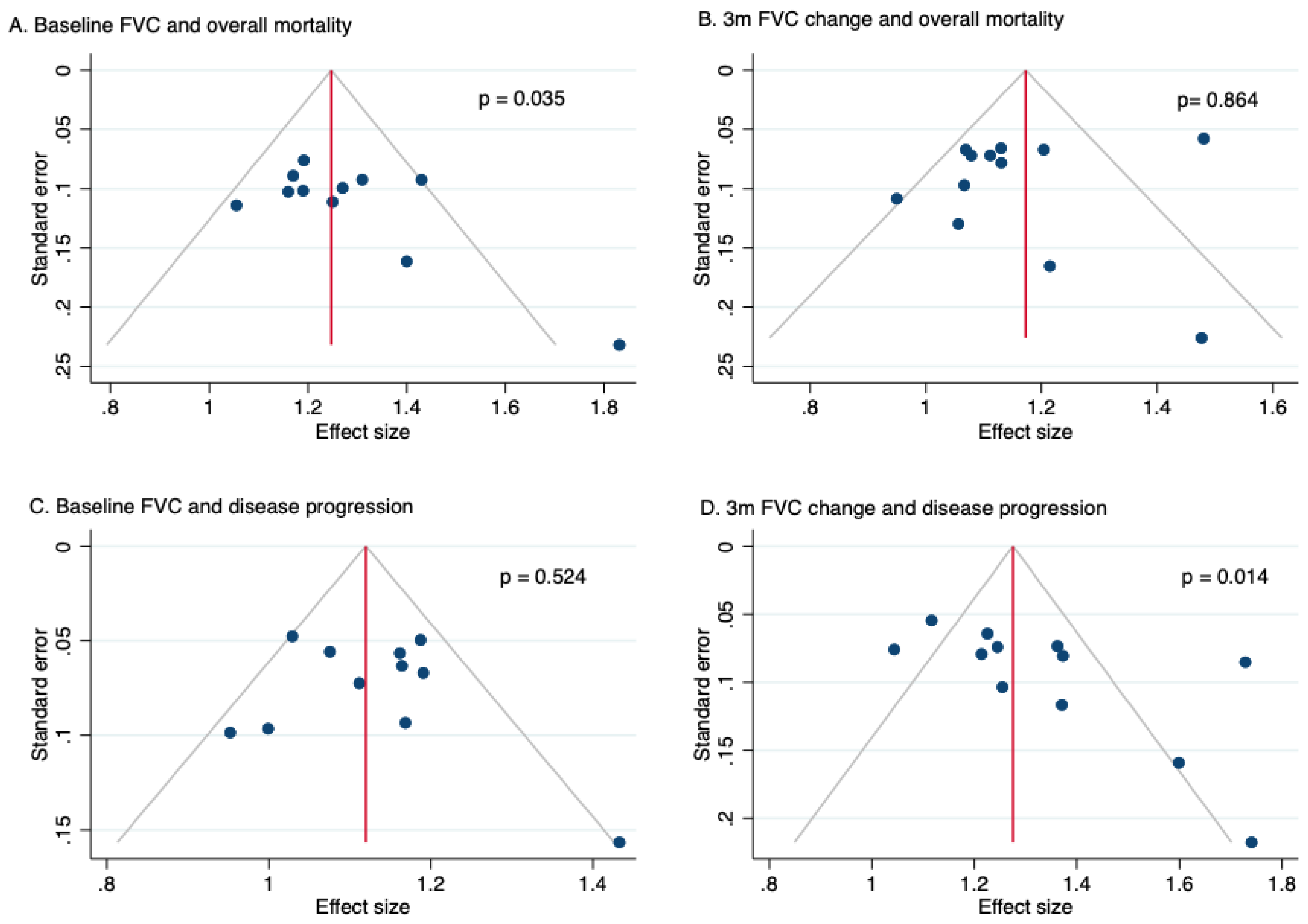


**Figure E11** – FVC publication bias. Publication bias assessed using Egger's test where ≥10 studies were included.

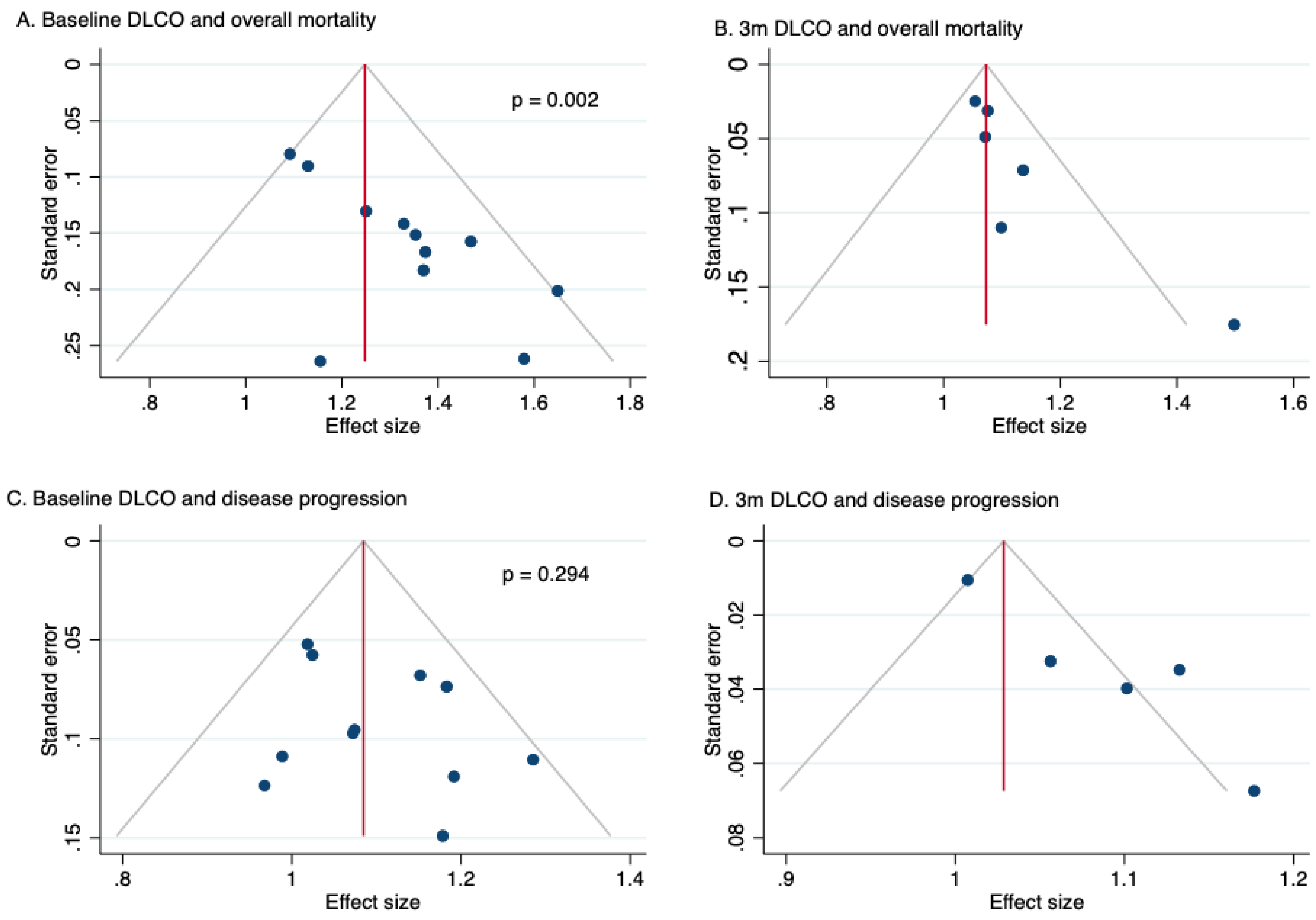


**Figure E12** – DLCO and publication bias. Publication bias assessed using Egger's test where ≥10 studies were included.

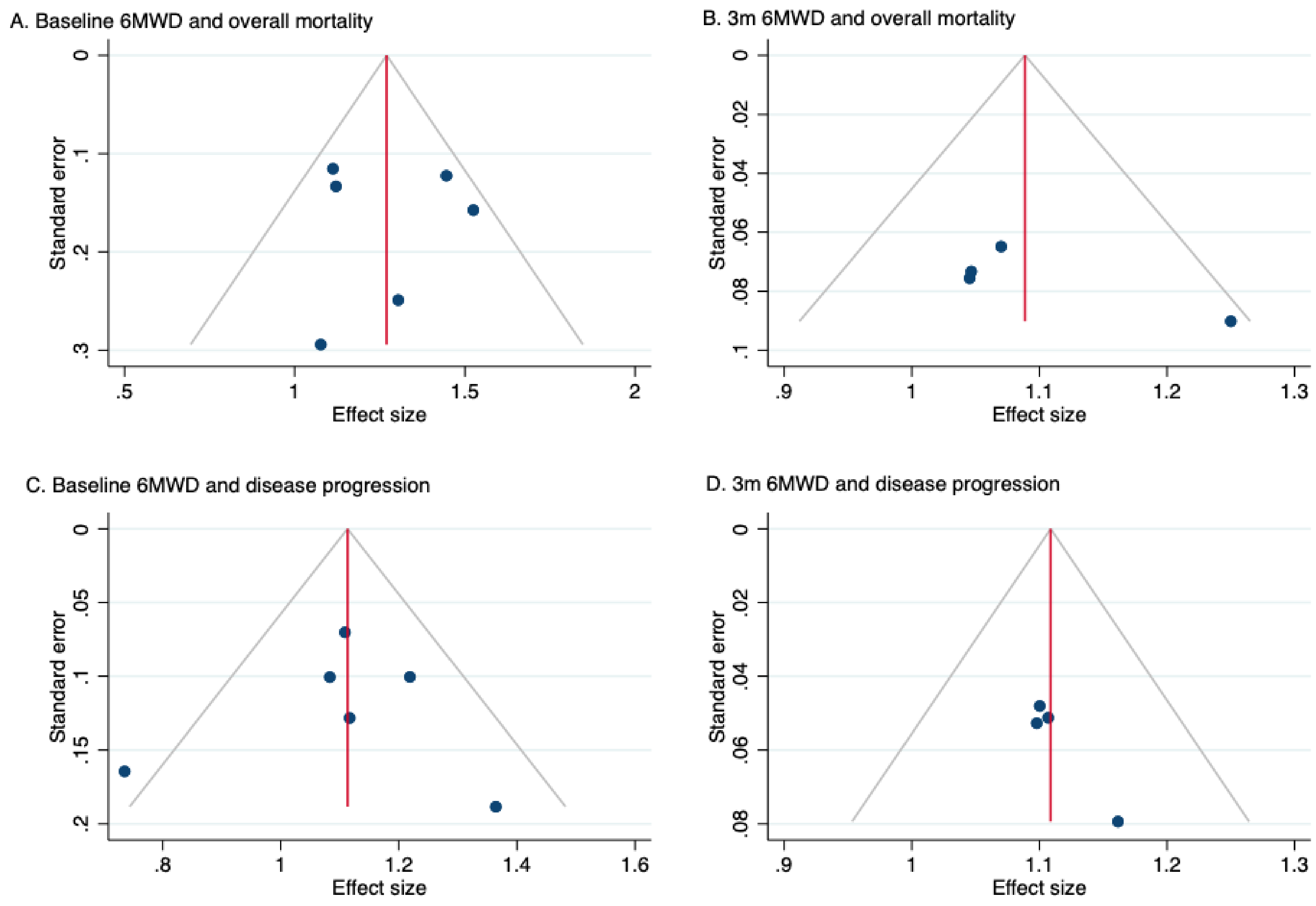


**Figure E13** – 6MWD and publication bias. Publication bias assessed using Egger's test where ≥10 studies were included.

| Study | Study participation | Study attrition | Prognostic factor | Outcome | Confounding |
|---|---|---|---|---|---|
| ARTEMIS | Low | High | Moderate | Moderate | Low |
| ASCEND | Low | Low | Low | Low | Low |
| BUILD-1 | Low | Low | Moderate | Low | Low |
| BUILD-3 | Low | Low | Moderate | Low | Low |
| CAPACITY | Low | Low | Moderate | Low | Low |
| IFIGENIA | Low | Low | Moderate | Low | Low |
| INPULSIS | Low | Low | Moderate | Low | Low |
| MUSIC | Low | Low | Moderate | Low | Low |
| TIPAC | Low | Low | Moderate | Low | Low |
| TOMORROW | Low | Low | Low | Low | Low |

**Table E1** – Risk of bias assessment. The risk of bias across studies was rated as low, moderate or high risk in five categories using the modified QUIPs tool.

| | Baseline FVC | | | | | | 3-month change in FVC | | | | 3-month FVC threshold | | | | FVC threshold AUROC | | | |
|---|---|---|---|---|---|---|---|---|---|---|---|---|---|---|---|---|---|---|
| Variables | Overall mortality | | Disease progression | | Change in FVC over 12m | | Overall mortality | | Disease progression | | Overall mortality (>5.7% decline) | | Disease progression (3% decline) | | Overall mortality | | Disease progression | |
| | $R^2$ (%) | P value | $R^2$ (%) | P value | $R^2$ (%) | P value | $R^2$ (%) | P value | $R^2$ (%) | P value | $R^2$ (%) | P value | $R^2$ (%) | P value | $R^2$ (%) | P value | $R^2$ (%) | P value |
| FVC vs. non GLI | 0.00 | 0.199 | 0.00 | 0.774 | 17.79 | 0.191 | 0.00 | 0.995 | 0.00 | 0.947 | 8.15 | 0.163 | 0.00 | 0.260 | 2.84 | 0.297 | 6.37 | 0.175 |
| Concomitant steroid use | 0.00 | 0.413 | 0.00 | 0.269 | 0.00 | 0.381 | 2.14 | 0.343 | **31.65** | **0.036** | 0.00 | 0.838 | 16.67 | 0.119 | 0.00 | 0.584 | **34.74** | **0.041** |
| Inclusion of severe cases | 0.00 | 0.628 | 0.00 | 0.811 | 22.21 | 0.157 | 0.00 | 0.927 | 5.99 | 0.304 | 0.00 | 0.497 | 0.00 | 0.327 | 0.00 | 0.644 | 0.00 | 0.411 |
| IPF diagnosis within 5 years | 4.94 | 0.475 | 0.00 | 0.811 | 0.00 | 0.84 | 0.00 | 0.859 | 0.00 | 0.476 | 0.00 | 0.871 | 0.00 | 0.476 | 0.00 | 0.622 | 0.00 | 0.473 |

| | Baseline $DL_{CO}$ | | | | | | 3-month change in $DL_{CO}$ | | | | 3-month $DL_{CO}$ threshold | | | | $DL_{CO}$ threshold AUROC | | | |
|---|---|---|---|---|---|---|---|---|---|---|---|---|---|---|---|---|---|---|
| Variables | Overall mortality | | Disease progression | | | | Overall mortality | | Disease progression | | Overall mortality (>10.5% decline) | | Disease progression (>7.2% decline) | | Overall mortality | | Disease progression | |
| | $R^2$ (%) | P value | $R^2$ (%) | P value | | | $R^2$ (%) | P value | $R^2$ (%) | P value | $R^2$ (%) | P value | $R^2$ (%) | P value | $R^2$ (%) | P value | $R^2$ (%) | P value |
| FVC vs. non GLI | 0.00 | 0.294 | 0.00 | 0.498 | | | 0.00 | 0.559 | **69.57** | **0.025** | 100 | 0.092 | 0.00 | 0.770 | 57.99 | 0.062 | 0.00 | 0.805 |
| Concomitant steroid use | **94.17** | **0.005** | 99.99 | 0.077 | | | 98.96 | 0.575 | 0.00 | 0.785 | 100 | 0.310 | 0.00 | 0.810 | **75.13** | **0.017** | 0.00 | 0.720 |
| Inclusion of severe cases | 0.00 | 0.681 | 0.00 | 0.686 | | | 0.00 | 0.927 | 0.00 | 0.787 | 100 | 0.180 | 0.00 | 0.77 | **100** | **0.000** | 0.00 | 0.805 |
| IPF diagnosis within 5 years | 0.00 | 0.681 | 0.00 | 0.811 | | | 0.00 | 0.859 | 0.00 | 0.787 | N/A | N/A | 50.63 | 0.088 | **100** | **0.000** | 26.22 | 0.233 |

| | Baseline 6MWD | | | | | | 3-month change in 6MWD | | | |
|---|---|---|---|---|---|---|---|---|---|---|
| Variables | Overall mortality | | Disease progression | | | | Overall mortality | | Disease progression | |
| | $R^2$ (%) | P value | $R^2$ (%) | P value | | | $R^2$ (%) | P value | $R^2$ (%) | P value |
| FVC vs. non GLI | N/A | N/A | N/A | N/A | | | N/A | N/A | N/A | N/A |
| Concomitant steroid use | **100** | **0.006** | 0.00 | 0.532 | | | **99.99** | **0.049** | 99.85 | 0.479 |
| Inclusion of severe cases | 0.00 | 0.479 | **65.17** | **0.017** | | | N/A | N/A | N/A | N/A |
| IPF diagnosis within 5 years | 0.00 | 0.916 | **65.17** | **0.017** | | | N/A | N/A | N/A | N/A |

**Table E2 -** Results of meta-regression for variables assessed separated by study outcomes. Sample sizes for each outcome shown (n). $R^2$ and p values from meta-regression shown where applicable. Significant associations (p<0.05) highlighted in red.
N/A, not applicable

| Outcome | The GRADE domains | Ratings for quality of evidence |
|---|---|---|
| **Forced Vital Capacity** | | |
| Overall mortality (11 studies, 1764 participants) | Risk of bias | All studies included well-defined participants diagnosed according to international consensus guidelines. Exposure and outcomes were measured objectively and consistently for all participants. FVC GLI corrections were applied. Covariates were adjusted for using IPD. Follow up was sufficiently long to enable outcomes to occur in all studies except one (ARTEMIS: terminated early due to futility) |
| | Imprecision | Effect sizes in all studies show a similar direction of effect. |
| | Inconsistency | No statistical heterogeneity as measured by $I^2$=0.0% |
| | Indirectness | No serious indirectness. All patients had IPF according to consensus criteria and were untreated. FVC was measured at baseline in all studies, and overall mortality measured from IPD. |
| | Publication bias | Publication bias present (p=0.035) |
| | Certainty of evidence | Moderate certainty of evidence |
| Disease progression (11 studies, 1526 participants) | Risk of bias | All studies included well-defined participants diagnosed according to international consensus guidelines. Exposure and outcomes were measured objectively and consistently for all participants. FVC GLI corrections were applied. Covariates were adjusted for using IPD. Follow up was sufficiently long to enable outcomes to occur in all studies except one (ARTEMIS: terminated early due to futility) |
| | Imprecision | Effect sizes in most studies show a similar direction of effect. |
| | Inconsistency | Low statistical heterogeneity as measured by $I^2$=25.9% |
| | Indirectness | No serious indirectness. All patients had IPF according to consensus criteria and were untreated. FVC was measured at baseline in all studies, and disease progression definition standardised using IPD. |
| | Publication bias | No evidence of publication bias |
| | Certainty of evidence | High certainty of evidence |

| Change in FVC over 3 months | | |
|---|---|---|
| Overall mortality (12 studies, 1729 participants) | Risk of bias | All studies included well-defined participants diagnosed according to international consensus guidelines. Exposure and outcomes were measured objectively and consistently for all participants. FVC GLI corrections were applied. Covariates were adjusted for using IPD. Follow up was sufficiently long to enable outcomes to occur in all studies except one (ARTEMIS: terminated early due to futility) |
| | Imprecision | Effect sizes in all studies show a similar direction of effect |
| | Inconsistency | Statistical heterogeneity as measured by $I^2$=59.4% |
| | Indirectness | No serious indirectness. All patients had IPF according to consensus criteria and were untreated. FVC was measured at baseline and three months in all included studies, and overall mortality measured from IPD. |
| | Publication bias | No evidence of publication bias |
| | Certainty of evidence | Moderate certainty of evidence |
| Disease progression (12 studies, 1551 participants) | Risk of bias | All studies included well-defined participants diagnosed according to international consensus guidelines. Exposure and outcomes were measured objectively and consistently for all participants. FVC GLI corrections were applied. Covariates were adjusted for using IPD. Follow up was sufficiently long to enable outcomes to occur in all studies except one (ARTEMIS: terminated early due to futility) |
| | Imprecision | Effect sizes in all studies show a similar direction of effect |
| | Inconsistency | Substantial heterogeneity ($I^2$=66.1%) |
| | Indirectness | No serious indirectness. All patients had IPF according to consensus criteria and were untreated. FVC was measured at baseline and three months in all included studies and disease progression standardised using IPD. |
| | Publication bias | Publication bias present (p=0.014) |
| | Certainty of evidence | Moderate certainty of evidence. |

| **$DL_{CO}$** | | |
|---|---|---|
| Overall mortality (11 studies; 1734 participants) | Risk of bias | All studies included well-defined participants diagnosed according to international consensus guidelines. Exposure and outcomes were measured objectively and consistently for all participants. Covariates were adjusted for using IPD. Follow up was sufficiently long to enable outcomes to occur in all studies except one (ARTEMIS: terminated early due to futility) |
| | Imprecision | Effect sizes in all studies showed a similar direction of effect with varying magnitude |
| | Inconsistency | No statistical heterogeneity as measured by $I^2$=0.0% |
| | Indirectness | No serious indirectness. All patients had IPF according to consensus criteria and were untreated. DLCO was measured at baseline in all included studies, and overall mortality measured from IPD. |
| | Publication bias | Publication bias present (p=0.002) |
| | Certainty of evidence | Moderate certainty of evidence |
| Disease progression (11 studies; 1512 participants) | Risk of bias | All studies included well-defined participants diagnosed according to international consensus guidelines. Exposure and outcomes were measured objectively and consistently for all participants. Covariates were adjusted for using IPD. Follow up was sufficiently long to enable outcomes to occur in all studies except one (ARTEMIS: terminated early due to futility) |
| | Imprecision | Effect sizes in all studies showed a similar direction of effect with varying magnitude |
| | Inconsistency | No statistical heterogeneity as measured by $I^2$=0.0% |
| | Indirectness | No serious indirectness. All patients had IPF according to consensus criteria and were untreated. DLCO was measured at baseline in all included studies, and disease progression definition standardised using IPD. |
| | Publication bias | No publication bias present |
| | Certainty of evidence | High certainty of evidence |

| **Change in $DL_{CO}$ over 3 months** | | |
|---|---|---|
| Overall mortality (6 studies, 736 participants) | Risk of bias | All studies included well-defined participants diagnosed according to international consensus guidelines. Exposure and outcomes were measured objectively and consistently for all participants. Covariates were adjusted for using IPD. Follow up was sufficiently long to enable outcomes to occur in all studies. $DL_{CO}$ at 3 months was only available from 6/11 datasets introducing a possible bias |
| | Imprecision | Effect sizes in all studies show a similar magnitude and direction of effect |
| | Inconsistency | No statistical heterogeneity as measured by $I^2$=0.0% |
| | Indirectness | No serious indirectness. All patients had IPF according to consensus criteria and were untreated. DLCO was measured at baseline and three months in all included studies, and overall mortality measured from IPD. |
| | Publication bias | No obvious funnel plot asymmetry. Unable to perform Egger's test due to fewer than 10 studies |
| | Certainty of evidence | Moderate certainty of evidence |
| Disease progression (12 studies, 1551 participants) | Risk of bias | All studies included well-defined participants diagnosed according to international consensus guidelines. Exposure and outcomes were measured objectively and consistently for all participants. Covariates were adjusted for using IPD. Follow up was sufficiently long to enable outcomes to occur in all studies. $DL_{CO}$ at 3 months was only available from 5/11 datasets introducing a possible bias |
| | Imprecision | Effect sizes in all studies show a similar direction of effect |
| | Inconsistency | Substantial heterogeneity ($I^2$=79.2%) |
| | Indirectness | No serious indirectness. All patients had IPF according to consensus criteria and were untreated. DLCO was measured at baseline and three months in all included studies and disease progression standardised using IPD. |
| | Publication bias | Funnel plot asymmetry but unable to perform Egger's test due to fewer than 10 studies |

| | | |
|---|---|---|
| | Certainty of evidence | Moderate certainty of evidence. |
| **Baseline 6MWD** | | |
| Overall mortality (6 studies, 828 participants) | Risk of bias | All studies included well-defined participants diagnosed according to international consensus guidelines. Exposure and outcomes were measured objectively and consistently for all participants. Covariates were adjusted for using IPD. Follow up was sufficiently long to enable outcomes to occur in all studies except one (ARTEMIS: terminated early due to futility) |
| | Imprecision | Effect sizes in all studies show a similar magnitude and direction of effect |
| | Inconsistency | Low heterogeneity ($I^2$=0.0%) |
| | Indirectness | No serious indirectness. All patients had IPF according to consensus criteria and were untreated. 6MWD was measured at baseline in all included studies and overall mortality measured from IPD. |
| | Publication bias | No obvious funnel plot asymmetry. Unable to perform Egger's test due to fewer than 10 studies |
| | Certainty of evidence | High certainty of evidence. |
| Disease progression (5 studies, 718 participants) | Risk of bias | All studies included well-defined participants diagnosed according to international consensus guidelines. Exposure and outcomes were measured objectively and consistently for all participants. Covariates were adjusted for using IPD. Follow up was sufficiently long to enable outcomes to occur in all studies except one (ARTEMIS: terminated early due to futility) |
| | Imprecision | Effect sizes in all studies show a similar magnitude and direction of effect, though overall estimate is inconclusive. |
| | Inconsistency | Moderate heterogeneity ($I^2$=40.4%) |
| | Indirectness | No serious indirectness. All patients had IPF according to consensus criteria and were untreated. 6MWD was measured at baseline and three months in all included studies and disease progression standardised using IPD. |
| | Publication bias | No obvious funnel plot asymmetry. Unable to perform Egger's test due to fewer than 10 studies |

| | | |
|---|---|---|
| | Certainty of evidence | Low certainty of evidence |
| **Change in 6MWD over 3 months** | | |
| Overall mortality (4 studies, 696 participants) | Risk of bias | All studies included well-defined participants diagnosed according to international consensus guidelines. Exposure and outcomes were measured objectively and consistently for all participants. Covariates were adjusted for using IPD. Follow up was sufficiently long to enable outcomes to occur. 3-month 6MWD available from 4/6 of datasets. |
| | Imprecision | Effect sizes in all studies show a similar magnitude and direction of effect |
| | Inconsistency | No statistical heterogeneity as measured by $I^2$=0.0% |
| | Indirectness | No serious indirectness. All patients had IPF according to consensus criteria and were untreated. $DL_{CO}$ was measured at baseline and three months in all included studies, and overall mortality measured from IPD. |
| | Publication bias | Too few studies to assess |
| | Certainty of evidence | High certainty of evidence |
| Disease progression (4 studies, 691 participants) | Risk of bias | All studies included well-defined participants diagnosed according to international consensus guidelines. Exposure and outcomes were measured objectively and consistently for all participants. Covariates were adjusted for using IPD. Follow up was sufficiently long to enable outcomes to occur. 3-month 6MWD available from 4/6 of datasets. |
| | Imprecision | Effect sizes in all studies show a similar direction of effect |
| | Inconsistency | High heterogeneity ($I^2$=79.2%) |
| | Indirectness | No serious indirectness. All patients had IPF according to consensus criteria and were untreated. DLCO was measured at baseline and three months in all included studies and disease progression standardised using IPD. |
| | Publication bias | Too few studies to assess |

| | | |
|---|---|---|
| | Certainty of evidence | Moderate certainty of evidence. |
| **3-month thresholds (FVC)** | | |
| Pooled threshold for mortality | Risk of bias | All studies included well-defined participants diagnosed according to international consensus guidelines. Exposure and outcomes were measured objectively and consistently for all participants. Covariates were adjusted for using IPD. Follow up was sufficiently long to enable outcomes to occur in all studies except one (ARTEMIS: terminated early due to futility) |
| | Imprecision | Optimal thresholds showed a similar direction of effect |
| | Inconsistency | No heterogeneity detected ($I^2$ = 0%) |
| | Indirectness | No serious indirectness. All patients had IPF according to consensus criteria and were untreated. FVC was measured at baseline and three months in all included studies, and overall mortality measured from IPD. |
| | Publication bias | Not applicable |
| | Certainty of evidence | High certainty of evidence |
| Pooled threshold for disease progression | Risk of bias | All studies included well-defined participants diagnosed according to international consensus guidelines. Exposure and outcomes were measured objectively and consistently for all participants. Covariates were adjusted for using IPD. Follow up was sufficiently long to enable outcomes to occur in all studies except one (ARTEMIS: terminated early due to futility) |
| | Imprecision | Optimal thresholds showed a similar direction of effect in all studies except one |
| | Inconsistency | Low heterogeneity detected ($I^2$ = 31.3%) |
| | Indirectness | No serious indirectness. All patients had IPF according to consensus criteria and were untreated. FVC was measured at baseline and three months in all included studies, and disease progression standardised and measured from IPD. |
| | Publication bias | Not applicable |

| | | |
|---|---|---|
| | Certainty of evidence | High certainty of evidence |
| Summary estimates for mortality using 3-month threshold | Risk of bias | All studies included well-defined participants diagnosed according to international consensus guidelines. Exposure and outcomes were measured objectively and consistently for all participants. Covariates were adjusted for using IPD. Follow up was sufficiently long to enable outcomes to occur in all studies except one (ARTEMIS: terminated early due to futility) |
| | Imprecision | Effect sizes in all studies show a similar direction of effect |
| | Inconsistency | Low heterogeneity ($I^2$=25.2%) |
| | Indirectness | No serious indirectness. All patients had IPF according to consensus criteria and were untreated. FVC was measured at baseline and three months in all included studies, and mortality measured from IPD. |
| | Publication bias | Not applicable |
| | Certainty of evidence | High certainty of evidence |
| Summary estimates for disease progression using 3-month threshold | Risk of bias | All studies included well-defined participants diagnosed according to international consensus guidelines. Exposure and outcomes were measured objectively and consistently for all participants. Covariates were adjusted for using IPD. Follow up was sufficiently long to enable outcomes to occur in all studies except one (ARTEMIS: terminated early due to futility) |
| | Imprecision | Effect sizes in most studies show a similar direction of effect |
| | Inconsistency | Substantial heterogeneity ($I^2$=58.5%) |
| | Indirectness | No serious indirectness. All patients had IPF according to consensus criteria and were untreated. FVC was measured at baseline and three months in all included studies, and disease progression standardised from IPD. |
| | Publication bias | Not applicable |
| | Certainty of evidence | Moderate certainty of evidence |

| 3-month thresholds ($DL_{CO}$) | | |
|---|---|---|
| Pooled threshold for mortality | Risk of bias | All studies included well-defined participants diagnosed according to international consensus guidelines. Exposure and outcomes were measured objectively and consistently for all participants. Covariates were adjusted for using IPD. Follow up was sufficiently long to enable outcomes to occur in all studies. $DL_{CO}$ at 3 months was only available from 6/11 datasets introducing a possible bias |
| | Imprecision | Very wide confidence intervals in most summa estimates, and in pooled estimate |
| | Inconsistency | Low heterogeneity ($I^2$=19.9%) |
| | Indirectness | No serious indirectness. All patients had IPF according to consensus criteria and were untreated. $DL_{CO}$ was measured at baseline and three months in all included studies, and overall mortality measured from IPD. |
| | Publication bias | Not applicable |
| | Certainty of evidence | Low certainty of evidence |
| Pooled threshold disease progression | Risk of bias | All studies included well-defined participants diagnosed according to international consensus guidelines. Exposure and outcomes were measured objectively and consistently for all participants. Covariates were adjusted for using IPD. Follow up was sufficiently long to enable outcomes to occur in all studies. $DL_{CO}$ at 3 months was only available from 6/11 datasets introducing a possible bias |
| | Imprecision | Very wide confidence intervals in most summa estimates, and in pooled estimate |
| | Inconsistency | No heterogeneity detected (0.0%) |
| | Indirectness | No serious indirectness. All patients had IPF according to consensus criteria and were untreated. $DL_{CO}$ was measured at baseline and three months in all included studies, and disease progression standardised from IPD. |
| | Publication bias | Not applicable |

| | | |
|---|---|---|
| | Certainty of evidence | Low certainty of evidence |

**Table E3** – GRADE (Grading of Recommendations, Assessment, Development and Evaluations) approach to rate the quality of evidence for each of the physiological variables and outcomes

| Study and year | Reason for exclusion | Interventional drug | Study phase | Centre | Placebo sample size | Study follow up, months (median) | White ethnicity % | Age (years) | Sex – male (%) | Baseline FVC % predicted | Baseline $DL_{CO}$ % predicted | Baseline 6MWD |
|---|---|---|---|---|---|---|---|---|---|---|---|---|
| Azuma et al, 2005[1] | Data sharing not permitted by original ethics | Pirfenidone | Phase 2 | multi-centre | 36 | 9 | - | 64.3 (7.6) | 94 | 78.4 (17.2) | 57.7 (13.8) | - |
| Daniels et al, 2010[2] | Data sharing not permitted by original consent | Imatinib | Phase 2 | multi-centre | 61 | 22 | - | 67.8 (52-79) range | 64 | 65.6 (mean only) | 39.3 (mean only) | 379 |
| Homma et al, 2012[3] | Data sharing not permitted by original ethics | N-acetylcysteine | Phase 3 | multi-centre | 46 | 12 | - | 68.2 (7.7) | 76 | 88.7 (15.5) | 64.4 (20.1) | - |
| King et al, 2009 (INSPIRE)[4] | FDA restrictions on data sharing | Interferon Gamma-1b | Phase 3 | multi-centre | 275 | 18 | - | 65.9 (7.9) | 68 | 73.1 (13.4) | 47.3 (9.3) | 392.8 (112.9) |
| Malouf et al, 2011[5] | Data sharing not permitted by original ethics | Everolimus | Phase 2 | multi-centre | 45 | 36 | - | 60 (9) | 71 | 69 (20) | 42 (14) | 451 (118) |
| Martinez et al, 2014 (PANTHER)[6] | Data sharing not permitted by original ethics | N-Acetylcysteine /Azathioprine | Phase 3 | multi-centre | 131 | 14 | 96 | 67.2 (8.2) | 75 | 73.4 (14.3) | 46 (12.2) | 375 (105) |
| Noth et al 2012 (ACE-IPF)[7] | Data sharing not permitted by original ethics | Warfarin | Phase 3 | multi-centre | 73 | 12 | 93 | 66.7 (7.4) | 79 | 58.7 (16.1) | 34.6 (13.4) | 280.2 (136.2) |
| Palmer et al, 2018[8] | Offices closed due to Covid-19 | BMS-986020 | Phase 2 | multi-centre | 47 | 6 | 64 | 69 (49-85) range | 70 | 69 (48-96) range | 45 (12-73) range | - |
| Parker et al, 2018[9] | Data not yet submitted to regulatory authorities | Tralokinumab | Phase 2 | multi-centre | 59 | 16 | 75 | 67.5 (6.1) | 79 | 70.3 (12.0) | 47 (13.8) | 391 (112) |
| Raghu et al, 2004[10] | FDA restrictions on data sharing | Interferon Gamma-1b | Phase 3 | multi-centre | 168 | 13 | 86 | 63.4 (8.6) | 66 | 64.1 (11.3) | 36.8 (10.6) | - |
| Raghu et al, 2008[11] | Denied by sponsor | Etanercept | Phase 2 | multi-centre | 41 | 12 | - | 65.1 (7.1) | 59 | 63.0 (12.7) | 36.9 (10.8) | 396.8 (136.8) |
| Richeldi et al, 2020 (PRAISE)[12] | No response from study personnel | Pamrevlumab | Phase 2 | multi-centre | 53 | 12 | - | 68.4 (7.2) | 81 | 73.1 (11.1) | 53.8 (12.2) | - |
| Taniguchi et al, 2009[13] | Data sharing not permitted by original ethics | Pirfenidone | Phase 3 | multi-centre | 104 | 12 | - | 64.7 (7.3) | 78 | 79.1 (17.4) | 55.2 (18.2) | - |

**Table E4 -** Baseline participant characteristics from studies where IPD could not be retrieved.
Values for physiological variables reported in mean (standard deviation) unless otherwise stated. 6MWD, six-minute walk distance, $DL_{CO}$, gas transfer for carbon monoxide; FVC, forced vital capacity; MRC, medical research council; -, data not available

**References**


1. Azuma A, Nukiwa T, Tsuboi E, et al. Double-blind, placebo-controlled trial of pirfenidone in patients with idiopathic pulmonary fibrosis. *Am J Respir Crit Care Med* 2005;171(9):1040-7. doi: 10.1164/rccm.200404-571OC [published Online First: 2005/01/25]
2. Daniels CE, Lasky JA, Limper AH, et al. Imatinib treatment for idiopathic pulmonary fibrosis: Randomized placebo-controlled trial results. *Am J Respir Crit Care Med* 2010;181(6):604-10. doi: 10.1164/rccm.200906-0964OC [published Online First: 2009/12/17]
3. Homma S, Azuma A, Taniguchi H, et al. Efficacy of inhaled N-acetylcysteine monotherapy in patients with early stage idiopathic pulmonary fibrosis. *Respirology (Carlton, Vic)* 2012;17(3):467-77. doi: 10.1111/j.1440-1843.2012.02132.x [published Online First: 2012/01/20]
4. King TE, Jr., Albera C, Bradford WZ, et al. Effect of interferon gamma-1b on survival in patients with idiopathic pulmonary fibrosis (INSPIRE): a multicentre, randomised, placebo-controlled trial. *Lancet (London, England)* 2009;374(9685):222-8. doi: 10.1016/s0140-6736(09)60551-1 [published Online First: 2009/07/03]
5. Malouf MA, Hopkins P, Snell G, et al. An investigator-driven study of everolimus in surgical lung biopsy confirmed idiopathic pulmonary fibrosis. *Respirology (Carlton, Vic)* 2011;16(5):776-83. doi: 10.1111/j.1440-1843.2011.01955.x [published Online First: 2011/03/03]
6. Martinez FJ, de Andrade JA, Anstrom KJ, et al. Randomized trial of acetylcysteine in idiopathic pulmonary fibrosis. *The New England journal of medicine* 2014;370(22):2093-101. doi: 10.1056/NEJMoa1401739 [published Online First: 2014/05/20]
7. Noth I, Anstrom KJ, Calvert SB, et al. A placebo-controlled randomized trial of warfarin in idiopathic pulmonary fibrosis. *Am J Respir Crit Care Med* 2012;186(1):88-95. doi: 10.1164/rccm.201202-0314OC [published Online First: 2012/05/09]
8. Palmer SM, Snyder L, Todd JL, et al. Randomized, Double-Blind, Placebo-Controlled, Phase 2 Trial of BMS-986020, a Lysophosphatidic Acid Receptor Antagonist for the Treatment of Idiopathic Pulmonary Fibrosis. *Chest* 2018;154(5):1061-69. doi: 10.1016/j.chest.2018.08.1058 [published Online First: 2018/09/12]
9. Parker JM, Glaspole IN, Lancaster LH, et al. A Phase 2 Randomized Controlled Study of Tralokinumab in Subjects with Idiopathic Pulmonary Fibrosis. *Am J Respir Crit Care Med* 2018;197(1):94-103. doi: 10.1164/rccm.201704-0784OC [published Online First: 2017/08/09]
10. Raghu G, Brown KK, Bradford WZ, et al. A Placebo-Controlled Trial of Interferon Gamma-1b in Patients with Idiopathic Pulmonary Fibrosis. *New England Journal of Medicine* 2004;350(2):125-33. doi: 10.1056/NEJMoa030511
11. Raghu G, Brown KK, Costabel U, et al. Treatment of idiopathic pulmonary fibrosis with etanercept: an exploratory, placebo-controlled trial. *Am J Respir Crit Care Med* 2008;178(9):948-55. doi: 10.1164/rccm.200709-1446OC [published Online First: 2008/08/02]
12. Richeldi L, Fernández Pérez ER, Costabel U, et al. Pamrevlumab, an anti-connective tissue growth factor therapy, for idiopathic pulmonary fibrosis (PRAISE): a phase 2, randomised, double-blind, placebo-controlled trial. *The Lancet Respiratory medicine* 2020;8(1):25-33. doi: 10.1016/s2213-2600(19)30262-0 [published Online First: 2019/10/03]
13. Taniguchi H, Ebina M, Kondoh Y, et al. Pirfenidone in idiopathic pulmonary fibrosis. *The European respiratory journal* 2010;35(4):821-9. doi: 10.1183/09031936.00005209 [published Online First: 2009/12/10]
